# The effect of head motion on brain age prediction using deep convolutional neural networks

**DOI:** 10.1101/2023.11.03.23297761

**Authors:** Pál Vakli, Béla Weiss, Dorina Rozmann, György Erőss, Ádám Nárai, Petra Hermann, Zoltán Vidnyánszky

## Abstract

Deep learning can be used effectively to predict participants’ age from brain magnetic resonance imaging (MRI) data, and a growing body of evidence suggests that the difference between predicted and chronological age—referred to as brain-predicted age difference (brain-PAD)—is related to various neurological and neuropsychiatric disease states. A crucial aspect of the applicability of brain-PAD as a biomarker of individual brain health is whether and how brain-predicted age is affected by MR image artifacts commonly encountered in clinical settings. To investigate this issue, we trained and validated two different 3D convolutional neural network architectures (CNNs) from scratch and tested the models on a separate dataset consisting of motion-free and motion-corrupted T1-weighted MRI scans from the same participants, the quality of which were rated by neuroradiologists from a clinical diagnostic point of view. Our results revealed a systematic increase in brain-PAD with worsening image quality for both models. This effect was also observed for images that were deemed usable from a clinical perspective, with brains appearing older in medium than in good quality images. These findings were also supported by significant associations found between the brain-PAD and standard image quality metrics indicating larger brain-PAD for lower-quality images. Our results demonstrate a spurious effect of advanced brain aging as a result of head motion and underline the importance of controlling for image quality when using brain-predicted age based on structural neuroimaging data as a proxy measure for brain health.

**Highlights:**

- Two 3D CNNs trained from scratch and validated to predict age from T1-w brain MRI
- Testing on motion-free and motion-corrupted scans from the same participants
- Image quality assessed by neuroradiologists and using standard image quality metrics
- Systematic increase in brain-predicted age difference with worsening image quality
- Spurious advanced brain aging effect in scans deemed usable for clinical diagnostics

## 1. Introduction

The human brain undergoes considerable change in its structural architecture and functional organization during the adult lifespan. Age-related macrostructural changes are characterized by the expansion of the ventricular system and a heterogeneous pattern of reduction in gray matter volume [1–3], while changes in white matter volume follow an inverted U-shaped trajectory, with a peak around 50 years of age followed by a rapid decrease after 60 years of age [4,5]. Alterations in brain function are evidenced by the reorganization of the resting-state functional connectome [6] and changes in task-related brain activity across the lifespan [7]. Brain aging is also accompanied by cognitive decline—affecting in particular the speed of information processing and the faculties of reasoning, memory, and executive functions [8]—and an increased risk for neurodegenerative diseases such as Alzheimer’s disease and Parkinson disease [9]. However, it has also been shown that the pace and magnitude of structural changes in brain aging vary considerably among individuals [10,11]. Hence, quantifying the extent to which an individual deviates from the healthy brain aging trajectory allows for the identification of accelerated or decelerated brain aging and the evaluation of the associated risk for health deterioration and potential treatment options. To this end, the dominant approach in the last decade has been to predict the age of the individual in a machine learning framework based on structural or functional neuroimaging data. In the past few years, deep learning has been increasingly used to predict brain age from magnetic resonance imaging (MRI) data, in line with recent trends regarding the application of deep neural networks in medical image processing in general [12,13] and in the study of neurological and psychiatric disorders in particular [14,15]. In many cases, convolutional neural networks (CNNs) [16,17], designed to learn from raw data in the form of multi-dimensional arrays such as 2D images or 3D volumes [18] are used for this purpose. In particular, 3D CNNs have been used successfully to predict chronological age from minimally-preprocessed structural MRI scans [19–23].

A growing body of evidence suggests that the difference between the predicted brain age and the chronological age of the individual—referred to as brain-predicted age difference (brain-PAD)—is indicative of overall brain health [24,25], presenting a potential for its use as an aging biomarker. This difference has been shown to be smaller in individuals with autism spectrum disorder [26] and attention-deficit hyperactivity disorder [27] than in healthy control participants, in line with the assumption of delayed brain development in these patient populations. Conversely, having an older-appearing brain has been associated with an increased risk of mortality [28] and with various disease states including mild cognitive impairment and Alzheimer’s disease [29–31], epilepsy [32,33], and schizophrenia [34–36]. Longitudinal changes in brain age have also been associated with disease progression in dementia. Advanced brain aging has been related to cognitive decline and clinical disease severity, and increased brain-PAD has been demonstrated to accurately predict the conversion of mild cognitive impairment to Alzheimer’s disease [29,30]. This increase in brain-PAD has also been observed in cognitively unimpaired individuals that developed mild cognitive impairment or dementia later on [22]. Brain age prediction may also be applied effectively in the assessment of risk and severity in other neurological disease states. Evidence suggests that brain age is associated with cardiovascular risk factors including stroke risk [37], and disease severity and long-term recovery in post-stroke aphasia [38,39].

Given the potential of brain-predicted age as a biomarker of individual brain health, the feasibility and relevance of brain age prediction in clinical settings is an especially important issue. A recent study has shown that accurate brain age prediction can be achieved using clinical-grade T2-weighted MRI scans acquired in routine hospital examinations, with increased brain-PAD indicating excessive brain atrophy [21]. This suggests that high resolution scans optimized for research purposes may not be necessary to obtain diagnostically relevant information using brain age prediction. A related issue that remains to be investigated is whether and how brain-predicted age is affected by common MR image artifacts. One of the most prominent sources of degraded scan quality is patient motion, which can introduce ghosting and blurring artifacts in the MR image [40–43]. Head motion is a common problem in clinical MR examinations that entails significant costs to radiology departments [44]. Furthermore, when studying neurological and neuropsychiatric disorders or longitudinal changes in brain structure and function, head motion can be a confounding factor due to its correlation with age and disease state. Younger participants have been shown to move more during MR image acquisition than individuals aged between 20 and 40 years [45], and older adults are more prone to move their heads than younger ones [46]. Increased movement of the head is associated with autism spectrum disorder, attention-deficit hyperactivity disorder, and schizophrenia as well [45]. Head motion during functional magnetic resonance imaging (fMRI) has also been shown to differ systematically between patients with mild cognitive impairment, Alzheimer’s disease, and healthy controls [47]. The problem of motion is particularly significant because it is known to induce spurious effects in resting-state functional connectivity MRI [48–50] and structural brain MRI analysis [45,51]. A recent study has investigated the reliability and predictive power of three commonly used pre-trained brain age prediction algorithms—XGBoost, which uses gradient tree boosting to predict age based on precomputed morphometric features [52], and brainageR [28] and DeepBrainNet [53], using Gaussian process regression and 2D CNN, respectively, to predict age based on minimally preprocessed T1-weighted images—and observed a modest correlation between XGBoost brain-PAD and a combined measure of image quality derived using the Computational Anatomy Toolbox (CAT12) [54]. In the present work, we examined the sensitivity of 3D CNNs specifically to head motion. We investigated 3D CNNs due to their remarkable effectiveness in predicting brain age from structural and functional brain images [19–23]. We trained two different neural network architectures—3D CNN using spatially separable convolutions and a regression variant of the Simple Fully Convolutional Network (SFCN-reg) [20]—from scratch and validated them using a large number of T1-weighted MRI scans and tested the networks on the separate MR-ART dataset [55]. MR-ART consists of motion-free and motion-corrupted brain scans from the same participants, the quality of which were rated by neuroradiologists from a clinical point of view. In particular, we examined whether the degradation of image quality by motion artifacts has a significant effect on brain-predicted age, by comparing brain-PAD between different quality images of the same subjects. We focused on brain-PAD because any systematic motion-related effect may result in spurious differences when using brain-predicted age as a proxy for advanced or delayed brain aging. We also examined the relationship between brain-PAD and MRI image quality metrics (IQMs) that are known to be sensitive to MRI artifacts including those resulting from movement [56-59].

## 2. Methods

### 2.1. Data acquisition

#### 2.1.1. Source dataset

We used T1-weighted brain MRI scans from the UK Biobank dataset to train and evaluate the brain age prediction source model on a large cohort of predominantly healthy individuals with good quality images. UK Biobank is a large population-based prospective study with over 500,000 individuals recruited between 2006 and 2010 from across Great Britain and comprises detailed phenotypic and genotypic information about participants [60]. A subset of the participants underwent MRI assessment starting from 2014. Data were acquired on Siemens Skyra 3T MRI scanners (Siemens Healthcare, Erlangen, Germany), using a standard Siemens 32-channel RF receive head coil, at the UK Biobank imaging centers in Cheadle, Newcastle, and Reading. The brain imaging protocol included a T1-weighted 3D magnetization-prepared rapid gradient echo (MPRAGE) sequence for structural imaging, using in-plane acceleration (iPAT = 2) and a field-of-view (FOV) of 208 × 256 × 256 with isotropic 1 mm spatial resolution [61].

Image quality control on behalf of UK Biobank consisted of the rough manual review of T1 images supplemented by a beta-version automated quality control pipeline [61]. Participants with a T1-weighted brain MRI scan deemed “unusable” by the UK Biobank team were excluded from the present study. Images from participants who attended a second imaging visit were also discarded, as we plan to use this data in a further, unrelated analysis. In addition, participants with a self-reported history of cancer, stroke, heart attack, deep-vein thrombosis, or pulmonary embolism diagnosed by a medical doctor (based on data-fields 2453-2.0, 6150-2.0, and 6152-2.0) were also omitted from the current study. Eventually, data from N = 32897 participants (17371 females) were used. A single T1-weighted brain MRI scan was used from each subject to train and evaluate the brain age prediction model. The dataset was randomly divided into disjoint training (N = 26897), validation (N = 3000), and test sets (N = 3000) using stratified sampling with age and sex as stratification variables (Table 1.). Only data in the training and validation sets were used for model training and hyperparameter selection.

**Table 1.**
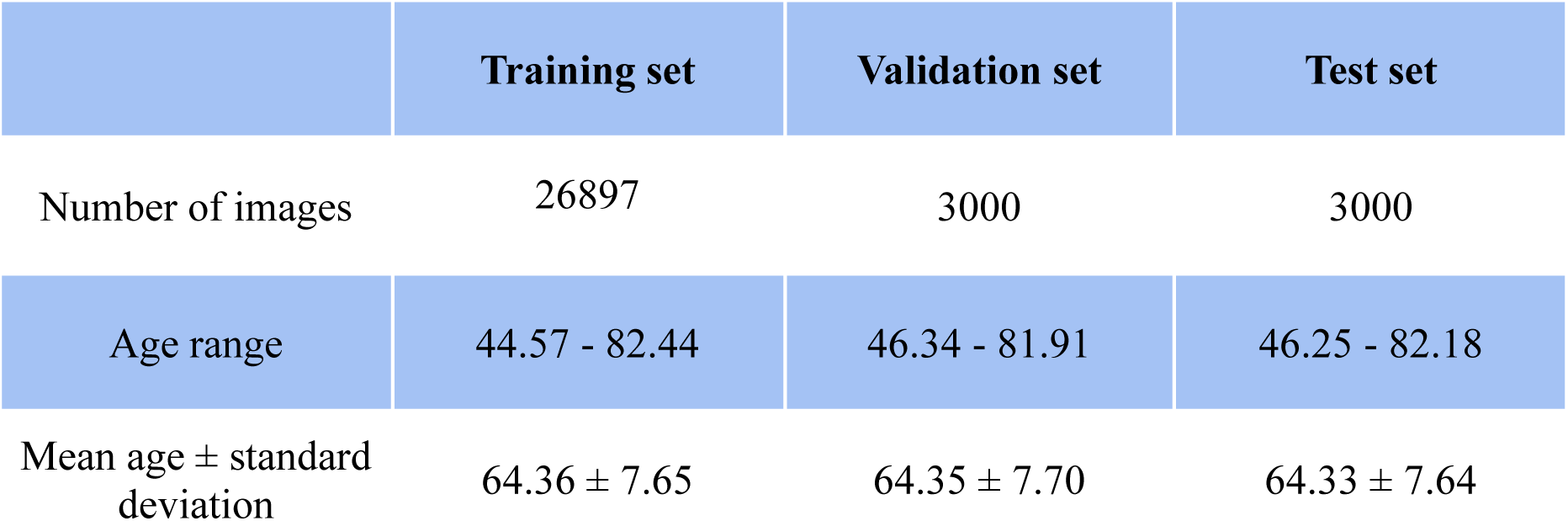
Number of images and descriptive statistics for chronological age (in years) at the time of the MRI assessment in the UK Biobank dataset used for the training and evaluation of the brain age prediction source model.

#### 2.1.2. Transfer learning dataset

The transfer learning dataset was used to adapt the source model to brain scans collected in our own lab. Importantly, it was ensured that there was no overlap in subjects between the transfer learning dataset and the MR-ART test set used to investigate the effect of head motion on brain age prediction (see section 2.1.3.). Image acquisitions were performed on a Siemens Magnetom Prisma 3T MRI scanner (Siemens Healthcare GmbH, Erlangen, Germany) with Siemens 20-channel, 32-channel, and 64-channel head-neck receiver coils at the Brain Imaging Centre, Research Centre for Natural Sciences. T1-weighted 3D MPRAGE anatomical images were acquired using 2-fold in-plane GRAPPA acceleration with isotropic 1 mm^3^ spatial resolution (repetition time (TR) = 2300 ms, echo time (TE) = 3 ms, inversion time (TI) = 900 ms, flip angle (FA) = 9°, FOV = 256 × 256 × 192 mm). The research protocol used for collecting the dataset was designed and conducted in accordance with the Hungarian regulations and laws, and was approved by the National Institute of Pharmacy and Nutrition (file number: OGYÉI/70184/2017).

The transfer learning dataset included 453 images from 451 participants (275 females). The dataset was divided into disjoint training (N = 361 images) and validation (N = 92) sets using stratification based on age, sex, and the number of head-neck receiver coil channels (Table 2.). The number of coils was taken into consideration because the MR-ART test set contained images acquired using the 20-channel head-neck receiver coil, however, we intended to use all available scans in our lab—apart from MR-ART images—to boost brain age prediction accuracy as much as possible.

**Table 2.** Number of images and descriptive statistics for chronological age (in years) at the time of image acquisition in the transfer learning dataset.

|  | Training set | Validation set |
| --- | --- | --- |
| Number of images<br>(number of subjects) | 361 (359) | 92 (92) |
| Age range | 18.14 - 89.48 | 18.35 - 79.95 |
| Mean age ± standard<br>deviation | 35.75 ± 18.82 | 36.23 ± 19.20 |

#### 2.1.3. MR-ART test set

We used MRI data collected in our own lab as part of the MR-ART dataset to investigate the accuracy of brain age prediction under different levels of image degradation related to head motion-induced artifacts. The MR-ART dataset [55] contains motion-free and motion-affected data acquired from the same participants. 148 healthy adult volunteers (95 females) with no reported history of neurological or psychiatric diseases participated in the study.

Image acquisitions were performed on a Siemens Magnetom Prisma 3T MRI scanner (Siemens Healthcare GmbH, Erlangen, Germany) with the standard Siemens 20-channel head-neck receiver coil at the Brain Imaging Centre, Research Centre for Natural Sciences. T1-weighted 3DMPRAGE anatomical images were acquired using 2-fold in-plane GRAPPA acceleration with isotropic 1 mm^3^ spatial resolution (TR = 2300 ms, TE = 3 ms, TI = 900 ms, FA = 9°, FOV = 256 × 256 mm). Three T1-weighted structural scans were acquired with the same parameters for each participant in a standard setting without motion (STAND) and with low (HM1) and high levels of head motion (HM2). During the acquisition, a fixation point was presented at the center of the display, and participants were instructed to gaze at this point. For the STAND scan, participants were instructed not to move at all, while for the HM1 and HM2 scans, participants were instructed to nod their head (tilt it down and then up along the sagittal plane) once every time the word “MOVE” (in Hungarian) appeared on the screen. To create different levels of movement artifacts, the word “MOVE” was presented 5 and 10 times evenly spaced during image acquisition for the HM1 and HM2 scans, respectively. Participants were instructed to avoid lifting their heads from the scanner table while nodding and to return their heads to the original position after performing a nod. Due to acquisition issues, one of the HM1 and HM2 scans is missing for 8 participants. All participants provided written, informed consent before participation. The research protocol used for collecting the dataset was designed and conducted in accordance with the Hungarian regulations and laws, and was approved by the National Institute of Pharmacy and Nutrition (file number: OGYÉI/70184/2017).

MR images were labeled based on visual inspection of the structural volumes by two neuroradiologists with more than ten years of experience, who were blind to the acquisition conditions (STAND, HM1 or HM2). Each record was rated on a 3-point scale based on image quality from the point of view of clinical diagnostic use. Clinically good (score 1), medium (score 2), and bad quality images (score 3) were differentiated, where bad quality images were considered unusable for clinical diagnostics (Table 3.). The two neuroradiologists initially harmonized their rating on 100 independent structural scans and were encouraged to discuss unclear cases during the whole labeling process in order to make the scores as robust as possible.

**Table 3.** Number of clinically good (score 1), medium (score 2), and bad quality (score 3) images and descriptive statistics for chronological age (in years) at the time of image acquisition in the MR-ART test set.

|  | Score 1 | Score 2 | Score 3 |
| --- | --- | --- | --- |
| Number of images<br>(number of subjects) | 129 (119) | 109 (82) | 198 (125) |
| Age range | 18.15 - 74.80 | 18.15 - 71.36 | 18.15 - 74.80 |
| Mean age ± standard deviation | 29.84 ± 12.18 | 29.51 ± 12.85 | 29.86 ± 12.24 |

### 2.2. Data preprocessing

Raw T1-weighted images comprising the source dataset were preprocessed by the UK Biobank team using an automated processing pipeline based on FSL tools [62]. The preprocessing pipeline included gradient distortion correction, cutting down the FOV, skull stripping, and non-linear transformation to MNI152 space [61]. The codes for the preprocessing pipeline are available online at: https://git.fmrib.ox.ac.uk/falmagro/UK_biobank_pipeline_v_1. We applied the same preprocessing pipeline to images in the transfer learning dataset and the MR-ART test set. Chronological age was calculated as the difference between the date of MR image acquisition and the date of birth divided by 365.25. As date of birth in the UK Biobank was available only with precision to the month, the 15th day of the month of birth was used to calculate age for each participant in the source dataset.

### 2.3. Neural network architectures

Two different 3D convolutional neural network architectures were used to perform brain age prediction based on volumetric T1-weighted MR images (Fig. 1). The first model, referred to as 3D CNN (Supplementary Table 1), was built using spatially separable convolutions to reduce the computational cost of processing whole-brain volumes. In this case, regular 3D convolutional layers are factorized into asymmetric convolutions—instead of convolving the input with kernels of shape K × K × K, a cascade of three convolutional operations with kernel shapes of K × 1 × 1, 1 × K × 1, and 1 × 1 × K, is applied to the images and feature maps. The proposed network consists of four convolutional blocks, with each block consisting of two sets of 3D spatially separable convolution, batch normalization [63] and rectified linear unit (ReLU) nonlinearity [64]. The first spatially separable convolution in the first convolutional block uses a stride of 2 to adaptively downsample the input volume, whereas all the remaining convolutional layers in the network use a stride of 1. Kernel size is 5 in the first and 3 in the remaining convolutional blocks, and all convolutional layers use SAME padding. The first two sets of spatially separable convolutions use 16 and 32 filters, and the number of filters is doubled in each successive block. Convolutional blocks are interleaved with max pooling layers to downsample feature maps at regular intervals. These pooling layers use a kernel size of 3, a stride of 2, and VALID padding. The last convolutional block is followed by global average pooling instead of max pooling. The spatial averages of feature maps in the last convolutional layer are fed into a fully connected hidden layer with 128 units using the ReLU nonlinearity, followed by a fully connected output layer consisting of a single unit with linear activation function. The overall neural network architecture is similar to the models applied in our previous studies for MRI quality control [65] and body mass index prediction based on T1-weighted MRI scans [66]. The model has 890,001 parameters, out of which 888,561 are trainable parameters.

**Fig. 1.**
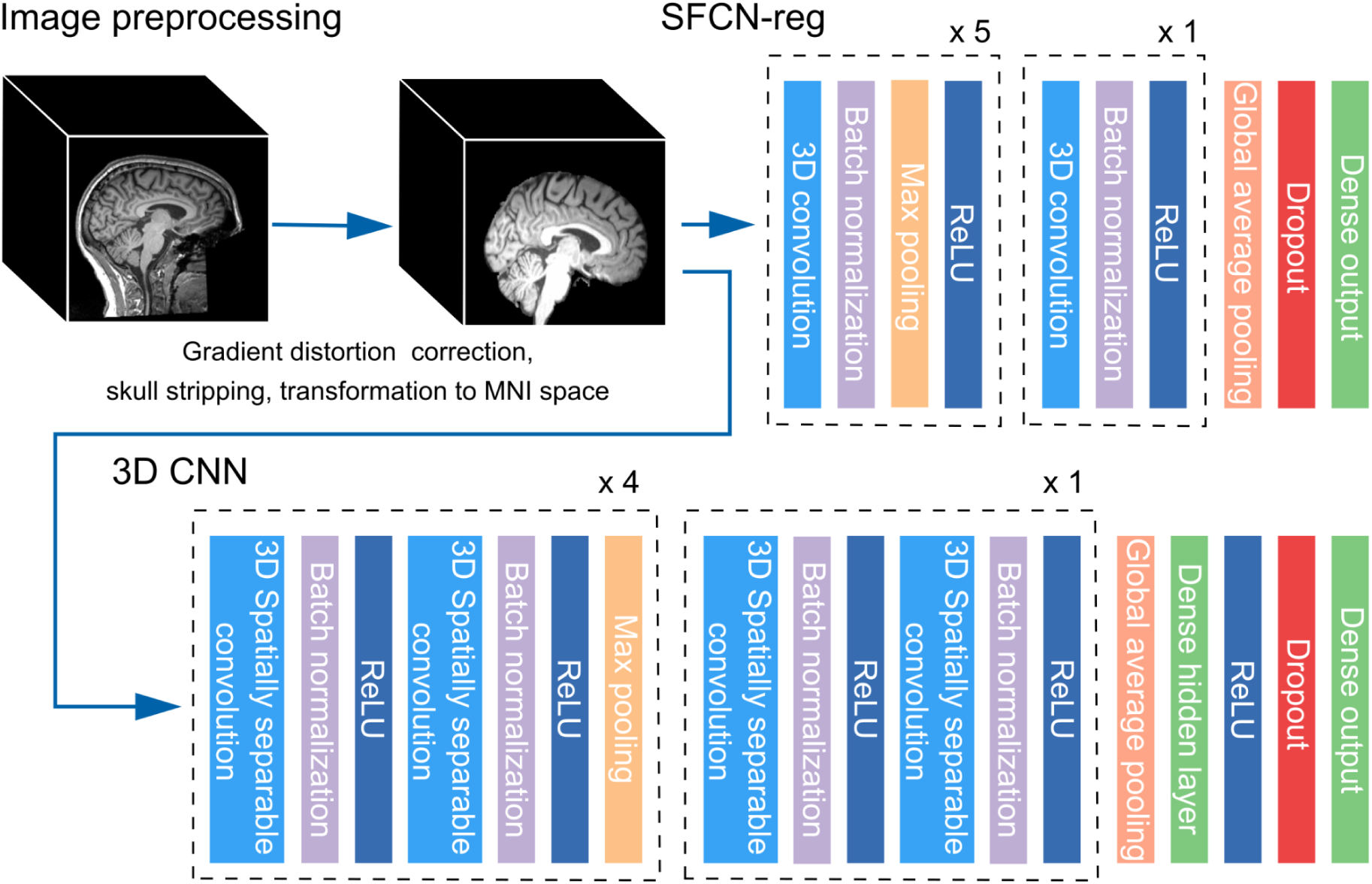
Overview of image preprocessing and neural network architectures (SFCN-reg and 3D CNN) used for brain age prediction. For a detailed description of the network layers, see Supplementary Table 1 and 2.

We also implemented a slightly modified version of the SFCN model developed by [20]. This network consists of 7 blocks. The first five blocks extract feature maps from the input image using regular 3D convolution, batch normalization, max pooling, and ReLU activation. The 6th block increases further the nonlinearity of the model by using pointwise convolution along with ReLU and batch normalization, without changing the size of the feature maps. The final block contains an average pooling layer and a fully connected layer along with softmax activation. The fully connected layer consists of 40 units, with the activation of each unit representing the probability that the participant’s age falls into a specific one-year age interval. In the currently implemented version of the model, referred to as SFCN-reg (Supplementary Table 2), the fully connected output layer consists of a single unit with no activation to represent brain-predicted age with a scalar value instead of a probability distribution. This implementation is similar to previous regression variants of the SFCN model [22,23].

The models were implemented in Python 3.8.13 using TensorFlow 2.4.1.

### 2.4. Training and evaluation

#### 2.4.1. Training and evaluating the source models

Both models were trained from scratch using the exact same UK Biobank training and validation set images (the source dataset). For the 3D CNN model, the weights of the convolutional layers and the fully connected hidden layer were initialized using He normal initialization [67] and the weights of the output layer were initialized using Glorot uniform initialization [68]. The bias terms of the convolutional and fully connected layers were initialized with zeros. We used the mean squared error (MSE) loss function and Adam optimization [69] with a learning rate of 0.0005 and a batch size of four. Dropout regularization [70] with a dropout rate of 0.4 was applied to the fully connected hidden layer during training. The network was trained using early stopping with a patience of 30 epochs, i.e. the model was evaluated on the validation set after each epoch and training was stopped when the validation loss had not decreased for the last 30 epochs.

For the SFCN-reg model, the weights of the layers were initialized using the default Glorot uniform initializer and the bias terms were initialized with zeros. The MSE loss was optimized using the Stochastic Gradient Descent (SGD) optimizer. Batch size was set to 4, and the learning rate was initialized to 0.01 and dropped by a factor of 0.3 every 30 epochs. L2 regularization was applied to the convolutional and fully connected layers with a regularization factor of 0.001, and dropout (with a dropout rate of 0.5) was applied after the global average pooling layer during training. The network was trained for 130 epochs, and model performance on the validation set was evaluated at the end of each epoch [20].

At the end of the training process, a snapshot of the model parameters leading to the best validation set performance (i.e. lowest MSE) was restored, and the models were evaluated on the UK Biobank test set to assess their generalizability. These models and the corresponding set of learnt weights constituted the source models that were fine-tuned using transfer learning (section 2.4.2.) and then evaluated on the MR-ART test set to assess their sensitivity to head motion-induced artifacts (2.4.3.). The accuracy of the models is estimated using the correlation coefficients between the predicted brain age and the chronological age (Pearson’s r and Spearman’s ρ), the mean absolute error (MAE), root mean squared error (RMSE), coefficient of determination (R^2^), and the mean and standard deviation of the brain-predicted age difference (brain-PAD, defined as predicted brain age minus chronological age). The age labels were divided by 100 during training and the model predictions were multiplied by 100 before the statistical analysis of the results.

A single NVIDIA RTX A6000 graphical processing unit (GPU) was used to train the networks.

#### 2.4.2. Transfer learning

The goal of transfer learning was to adapt the source models to MRI scans that were acquired using the same scanner and from participants in the same age range as in the MR-ART set. Again, it is important to note that there was no overlap in subjects between the transfer learning dataset and the MR-ART dataset. Transfer learning was performed in two phases, in a similar way for both models. In the first phase, the weights and bias term of the output layer were reinitialized and the network was trained for 10 epochs. The parameters of all layers other than the output layer were frozen during training, i.e. they were not allowed to be changed by backpropagation. The MSE loss was optimized using Adam for the 3D CNN and SGD for the SFCN-reg model. In the second phase, several layers below the output layer were unfreezed and the training process was continued using early stopping with a patience of 30 epochs. We experimented with unfreezing different numbers of layers and using different learning and dropout rates, and retained the configuration yielding the lowest validation MSE. Batch size was set to 4, and batch normalization layers were run in inference mode during the whole transfer learning process. The model parameters leading to the best transfer learning validation set performance were restored to evaluate the final models on the MR-ART dataset.

#### 2.4.3. Evaluation on the MR-ART test set

The final models were evaluated on the MR-ART test set to investigate the effects of head motion-induced artifacts on brain age prediction accuracy. Besides evaluating the models on the overall MR-ART set, we selected a subset of participants including all subjects with a clinically good quality (score 1) STAND, a medium quality (score 2) HM1, and a bad quality (score 3) HM2 image to ensure the independence of samples in each experimental condition and quality score level. We refer to this as image selection 1 - IS1. In this way, 105 images from 35 subjects (25 females; age range = 18.15 - 64.64 years; mean age ± standard deviation = 27.65 ± 9.78) were selected for further analysis. To compare brain-PAD across the three clinical scores/conditions, a two-way repeated measures analysis of variance (ANOVA) was performed with ‘clinical score’ (1/2/3) and ‘model’ (3D CNN/SFCN-reg) as within-subject factors. Greenhouse-Geisser corrected p-values are reported to account for violations of sphericity. Post-hoc analysis was performed using pairwise t-tests. Benjamini-Hochberg false discovery rate (FDR) correction was applied to account for the problem of multiple comparisons.

In a further step, we narrowed the analysis to good and medium quality images (image selection 2 - IS2). We selected all subjects who had a good quality (score 1) STAND image and at least one medium quality (score 2) image. If a subject had two medium quality images, the HM2 image was kept and used in the analysis. This way we were able to select good-medium quality image pairs from 54 subjects (36 females; age range = 18.15 - 71.36 years; mean age ± standard deviation = 29.57 ± 12.58). A two-way repeated measures ANOVA with ‘clinical score’ (1/2) and ‘model’ (3D CNN/SFCN-reg) as within-subject factors was performed to compare brain-PAD between good and medium quality images in these subjects. Post-hoc analysis was performed as detailed above. Metrics of model accuracy are also reported for each clinical score separately.

A particular bias is commonly observed in brain age prediction: the age of younger participants is overestimated and the age of elderly subjects is underestimated, whereas the age of participants whose age lies closer to the mean age in the training sample is estimated more accurately [71]. Besides the analyses reported in the manuscript using the original (uncorrected) model predictions, we rerun all the analyses twice after applying different correction procedures to address this potential bias. Importantly, we used the models’ predictions on the transfer learning validation set to perform these corrections (for details, see section S1.2 in Supplementary materials and methods).

Furthermore, to assess the association between image quality and brain-predicted age difference on the MR-ART dataset, partial Pearson’s and Spearman’s correlation coefficients were calculated between IQMs and brain-PAD by controlling for potential covariate effects of chronological age. The quality of images was characterized by the Euler number and some additional IQMs that were shown to be sensitive to MRI artifacts and head motion in particular [56-59]. The Euler numbers were obtained by running the FreeSurfer toolbox [72], and the following additional IQMs were selected from the MRIQC toolbox [58]: coefficient of joint variation (cjv), contrast-to-noise ratio (cnr), signal-to-noise ratio (snr, separately for cerebrospinal fluid, gray matter, white matter and total brain: snr_csf, snr_gm, snr_wm and snr_total), Dietrich’s SNR (snrd, similarly to SNR measures: snrd_csf, snrd_gm, snrd_wm and snrd_total) and quality indices QI1 and QI2. The MRIQC toolbox ran successfully on all MR-ART images, while FreeSurfer failed for a single image recorded in the HM2 condition and having clinical score 2. This image was excluded from correlations analysis. To control for performing multiple correlations, the obtained p values (p_u_) were corrected per correlation coefficient type (separately for Pearson’s and Spearman’s coefficients) by the Benjamini-Hochberg FDR method (p_c_). The IQMs were calculated using the MRIQC 0.16.1 and FreeSurfer 7.1.1 toolboxes. All statistical analyses were performed using the Pingouin 0.5.3. statistical package for Python 3 [73].

## 3. Results

### 3.1. Brain age prediction on the UK Biobank and the transfer learning datasets

3D CNN achieved particularly good age prediction performance on the UK Biobank validation set (MAE = 2.51 years, RMSE = 3.14 years, Pearson’s r = 0.91, Spearman’s ρ = 0.92, R^2^ = 0.83, mean brain-PAD ± standard deviation = 0.39 ± 3.11 years) and generalized very well to the UK Biobank test set (MAE = 2.55 years, RMSE = 3.20 years, Pearson’s r = 0.91, Spearman’s ρ = 0.91, R^2^ = 0.82, mean brain-PAD ± standard deviation = 0.44 ± 3.17 years). When applying transfer learning, the winning configuration consisted of fine-tuning the second set of spatially separable convolutions (starting with layer ‘conv_block_1/conv2_1’ in Supplementary Table 1) and all the layers above, using learning rates of 0.001 and 0.0001 in the first and second phase, respectively. This yielded a strong correlation between the brain-predicted and chronological age (Pearson’s r = 0.99, Spearman’s ρ = 0.81, p < 0.001 for both correlations) on the transfer learning validation set, and the model was highly accurate in predicting age (MAE = 2.63 years, RMSE = 3.20 years, R^2^ = 0.97, mean brain-PAD ± standard deviation = 0.12 ± 3.21 years; Fig. 2).

**Fig. 2.**
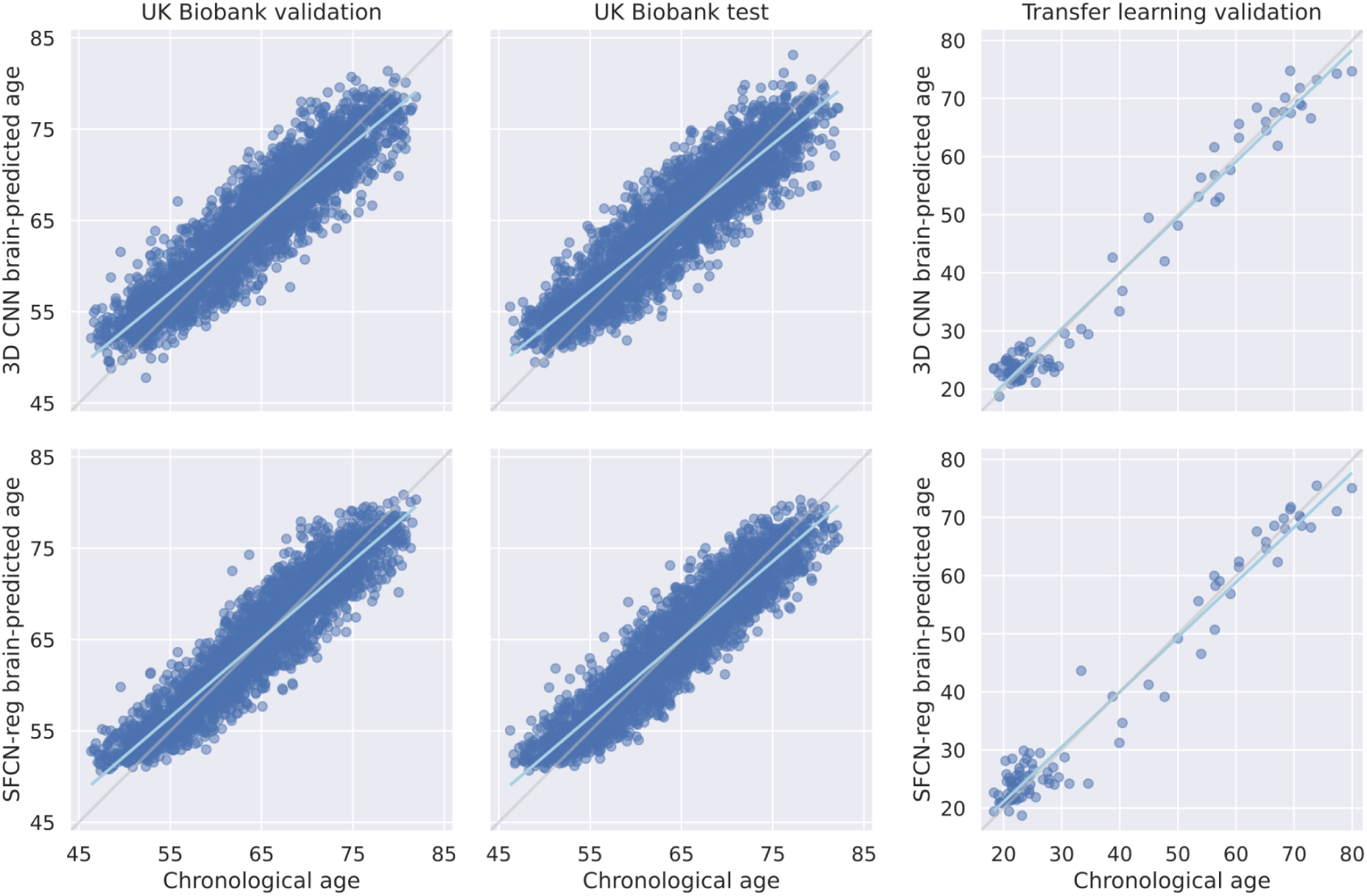
Correlation between chronological and brain-predicted age (in years) for the 3D CNN (upper row) and SFCN-reg (lower row) models on the UK Biobank validation and test sets, and the transfer learning validation set. Gray and cyan lines correspond to the lines of identity and the regression lines, respectively.

SFCN-reg achieved even better brain age prediction performance on the UK Biobank validation (MAE = 2.16 years, RMSE = 2.72 years, Pearson’s r = 0.94, Spearman’s ρ = 0.94, R^2^ = 0.88, mean brain-PAD ± standard deviation = 0.18 ± 2.71 years) and test sets (MAE = 2.17 years, RMSE = 2.71 years, Pearson’s r = 0.94, Spearman’s ρ = 0.94, R^2^ = 0.87, mean brain-PAD ± standard deviation = 0.14 ± 2.71 years) than the 3D CNN model. The winning transfer learning configuration consisted of fine-tuning the 6th convolutional layer (‘conv6’ in Supplementary Table 2) along with the reinitialized output layer, using a dropout rate of 0.1 and learning rates of 0.01 in the first and 0.001 with step decay in the second phase. This yielded remarkably good, albeit slightly lower brain age prediction accuracy on the transfer learning validation set (MAE = 2.92 years, RMSE = 3.73 years, Pearson’s r = 0.98, Spearman’s ρ = 0.85, R^2^ = 0.96, mean brain-PAD ± standard deviation = 0.19 ± 3.75 years; Fig. 2), than in the case of 3D CNN.

### 3.2. Brain-predicted age and clinical quality scores

Testing the fine-tuned models on the full MR-ART set (Table 4) revealed that the models generalized reasonably well in terms of absolute error to good quality (score 1; 3D CNN MAE = 3.67 years, SFCN-reg MAE = 3.20 years) and medium quality (score 2; 3D CNN MAE = 3.79 years, SFCN-reg MAE = 3.54 years) images. The results also show poor age prediction performance in the case of bad quality (score 3) images (Fig. 3), and an increase in brain-PAD with worsening image quality for both models (Fig. 4). For descriptive statistics and visualization of brain age prediction performance as a function of acquisition condition, see Supplementary Table 3 and Supplementary Fig. 1 and 2.

**Fig. 3.**
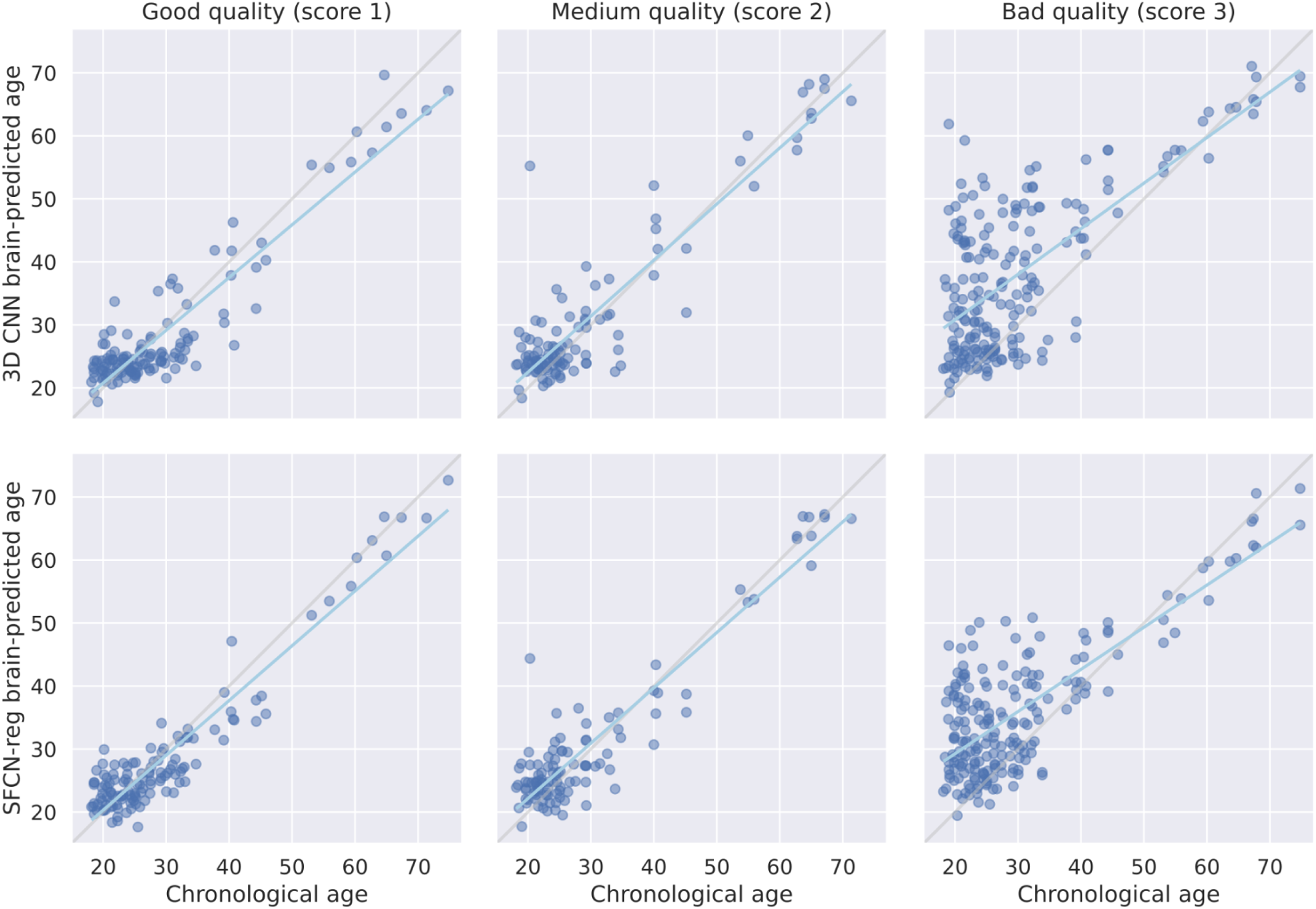
Correlations between chronological and brain-predicted age (in years) for the 3D CNN (upper row) and SFCN-reg (lower row) models on the full MR-ART set (N = 148 subjects) grouped according to image quality. Gray and cyan lines correspond to the lines of identity and the regression lines, respectively.

**Fig. 4.**
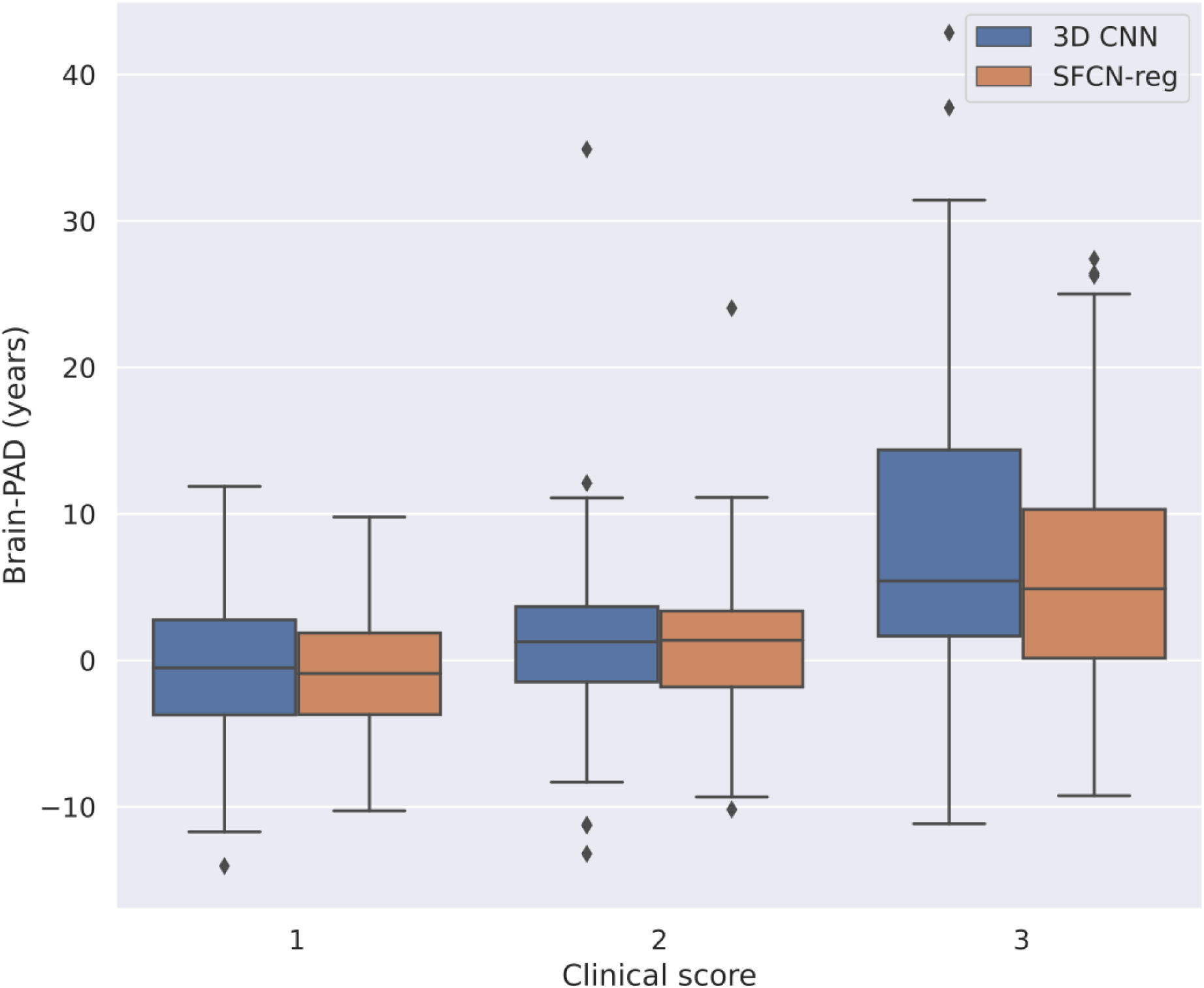
Distribution of brain-predicted age difference (brain-PAD) on the full MR-ART set (N = 148 subjects) for the clinically good (score 1), medium (score 2), and bad quality (score 3) images.

**Table 4.** Brain age prediction performance on the full MR-ART set (N = 148 subjects) for the clinically good (score 1), medium (score 2), and bad quality (score 3) images. All correlations are significant with p < 0.001. MAE = mean absolute error (in years), RMSE = root mean squared error (in years), R^2^ = coefficient of determination, brain-PAD = brain-predicted age difference (brain-predicted minus chronological age in years), std = standard deviation.

| Image quality | Good (N = 129) |  | Medium (N = 109) |  | Bad (N = 198) |  |
| --- | --- | --- | --- | --- | --- | --- |
| Model | 3D CNN | SFCN-reg | 3D CNN | SFCN-reg | 3D CNN | SFCN-reg |
| MAE | 3.67 | 3.20 | 3.79 | 3.54 | 9.35 | 7.42 |
| RMSE | 4.58 | 4.00 | 5.66 | 4.73 | 12.57 | 9.85 |
| $R^2$ | 0.85 | 0.89 | 0.81 | 0.87 | -0.10 | 0.32 |
| Pearson's r | 0.92 | 0.94 | 0.91 | 0.94 | 0.69 | 0.77 |
| Spearman's $\rho$ | 0.68 | 0.74 | 0.61 | 0.66 | 0.49 | 0.47 |
| mean brain-PAD ± std | -0.75 ± 4.54 | -0.91 ± 3.91 | 1.42 ± 5.50 | 0.97 ± 4.65 | 8.14 ± 9.60 | 6.09 ± 7.76 |

The results obtained when evaluating the fine-tuned models on the images of the subjects selected for ANOVA with clinical score as a three-level factor (IS1)—exhibited an overall similar pattern, with reasonably good prediction performance in terms of absolute error for good quality (score 1; 3D CNN MAE = 3.78 years, SFCN-reg MAE = 3.60 years) and medium quality (score 2; 3D CNN MAE = 3.58 years, SFCN-reg MAE = 3.59 years) images (see Table 5, Fig. 5 and 6). Brain-PAD was on average 2.70 years higher for medium (mean brain-PAD ± standard deviation = 1.62 ± 4.45 years) than good quality scans (mean brain-PAD ± standard deviation = -1.08 ± 4.42 years) for the 3D CNN model. A similar difference was observed for SFCN-reg, with brain-PAD being higher for medium (mean brain-PAD ± standard deviation = 0.92 ± 4.15 years) than good quality images (mean brain-PAD ± standard deviation = -1.33 ± 4.04 years). Brain age prediction performance was remarkably reduced in the case of bad quality (score 3) scans, with a substantial increase in mean absolute error (3D CNN MAE = 8.01 years, SFCN-reg MAE = 6.94 years) and brain-PAD (3D CNN mean brain-PAD ± standard deviation = 7.05 ± 7.98 years, SFCN-reg mean brain-PAD ± standard deviation = 5.79 ± 6.89 years) for both models. The repeated measures ANOVA revealed a significant difference in brain-PAD across clinical scores (F = 35.57, p < 0.001). There was no significant main effect of ‘model’ (F = 1.39, p = 0.2458), and the interaction was also not significant (F = 0.65, p = 0.4761). Post hoc analysis showed that brain-PAD significantly increased in the case of medium quality images when compared to good quality ones (p < 0.001). Brain-PAD also increased significantly for bad quality images when compared to either good (p < 0.001) or medium quality (p < 0.001) images.

**Fig. 5.**
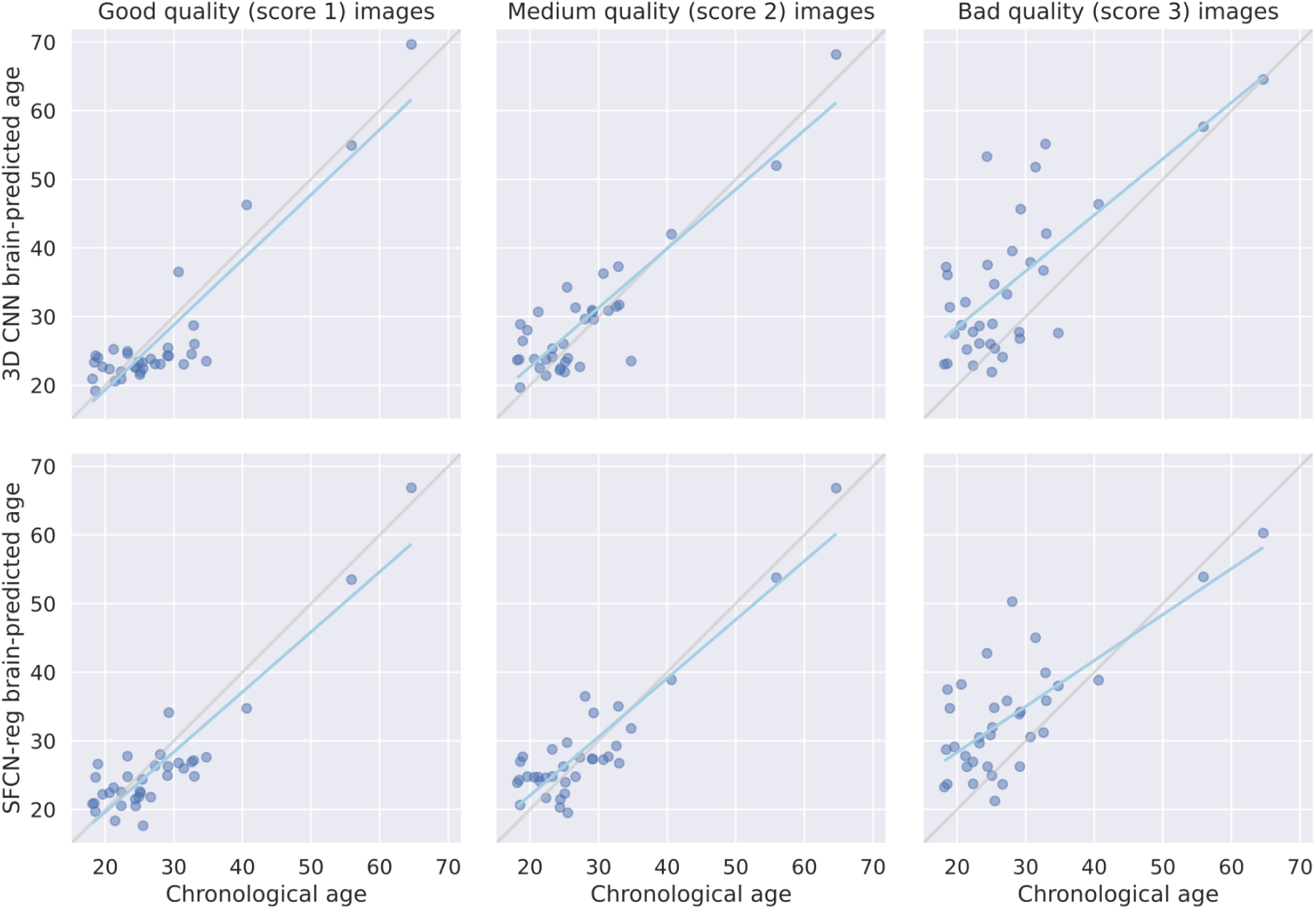
Correlations between chronological and brain-predicted age (in years) for the 3D CNN (upper row) and SFCN-reg (lower row) models for the selected MR-ART test set subjects (N = 35) with a clinically good (score 1), medium (score 2), and bad quality (score 3) image acquired without (STAND), with low (HM1), and with high (HM2) levels of head motion, respectively (IS1). Gray and cyan lines correspond to the lines of identity and the regression lines, respectively.

**Fig. 6.**
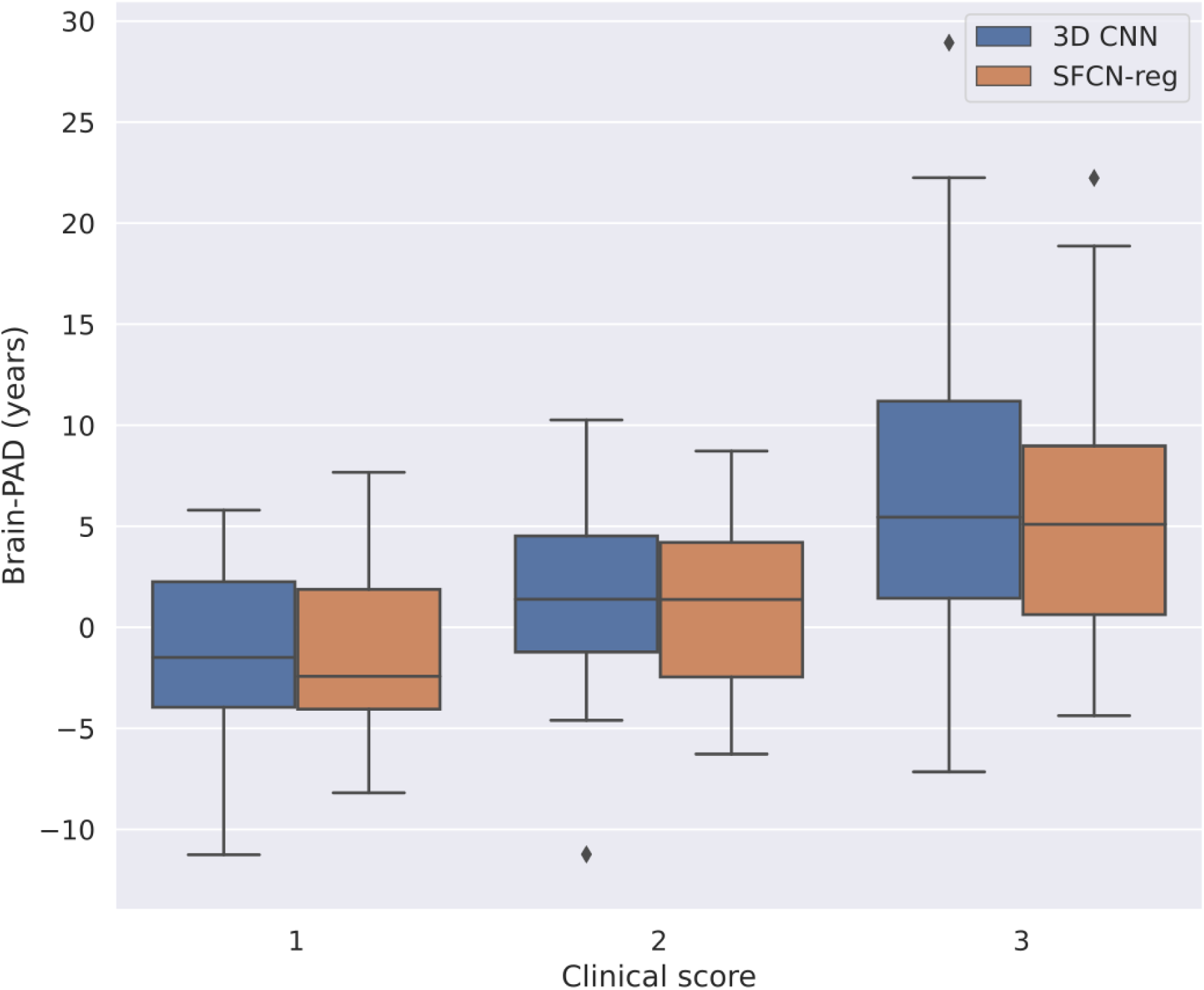
Distribution of brain-predicted age difference (brain-PAD) for the selected MR-ART test set subjects (N = 35) with a clinically good (score 1), medium (score 2), and bad quality (score 3) image acquired without (STAND), with low (HM1), and with high (HM2) levels of head motion, respectively (IS1). Brain-PAD is defined as brain-predicted minus chronological age in years.

**Table 5.** Brain age prediction performance for the selected MR-ART test set subjects (N = 35) with a clinically good (score 1), medium (score 2), and bad quality (score 3) image acquired without (STAND), with low (HM1), and with high (HM2) levels of head motion, respectively (IS1). All correlations are significant with p < 0.001. MAE = mean absolute error (in years), RMSE = root mean squared error (in years), R^2^ = coefficient of determination, brain-PAD = brain-predicted age difference (predicted minus chronological age in years), std = standard deviation.

| Image quality | Good ($N = 35$) | | Medium ($N = 35$) | | Bad ($N = 35$) | |
| --- | --- | --- | --- | --- | --- | --- |
| Model | 3D CNN | SFCN-reg | 3D CNN | SFCN-reg | 3D CNN | SFCN-reg |
| MAE | 3.78 | 3.60 | 3.58 | 3.59 | 8.01 | 6.94 |
| RMSE | 4.49 | 4.20 | 4.68 | 4.19 | 10.56 | 8.93 |
| $R^2$ | 0.78 | 0.81 | 0.76 | 0.81 | -0.20 | 0.14 |
| Pearson's $r$ | 0.90 | 0.91 | 0.89 | 0.91 | 0.72 | 0.73 |
| Spearman's $\rho$ | 0.60 | 0.68 | 0.63 | 0.66 | 0.53 | 0.53 |
| mean brain-PAD ± std | -1.08 ± 4.42 | -1.33 ± 4.04 | 1.62 ± 4.45 | 0.92 ± 4.15 | 7.05 ± 7.98 | 5.79 ± 6.89 |

With regard to the analysis focusing on the differences between good and medium quality images using all available score 1-score 2 image pairs (IS2;), a significant difference in brain-PAD was also observed in this case, with brain-PAD being higher for medium than good quality images (main effect of ‘clinical score’: F = 31.45, p < 0.001; Table 6, Fig. 7 and 8) There was no significant main effect of ‘model’ (F = 0.16, p = 0.6899), and the interaction was also not significant (F = 0.06, p = 0.8108).

**Fig. 7.**
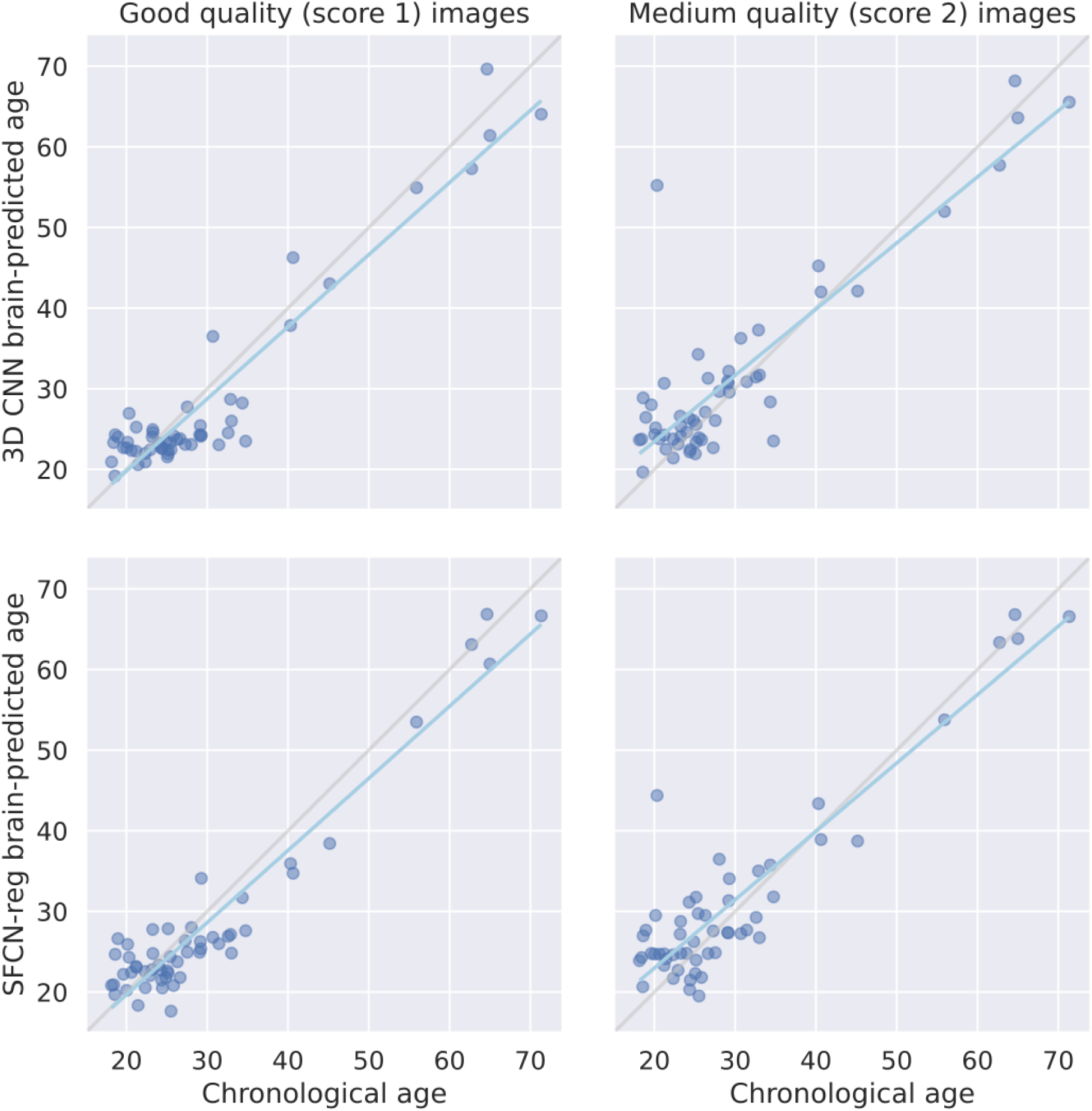
Correlations between chronological and brain-predicted age (in years) for the 3D CNN (upper row) and SFCN-reg (lower row) models for all MR-ART subjects (N = 54) with a good (score 1) - medium (score 2) quality image pair (IS2). Gray and cyan lines correspond to the lines of identity and the regression lines, respectively.

**Fig. 8.**
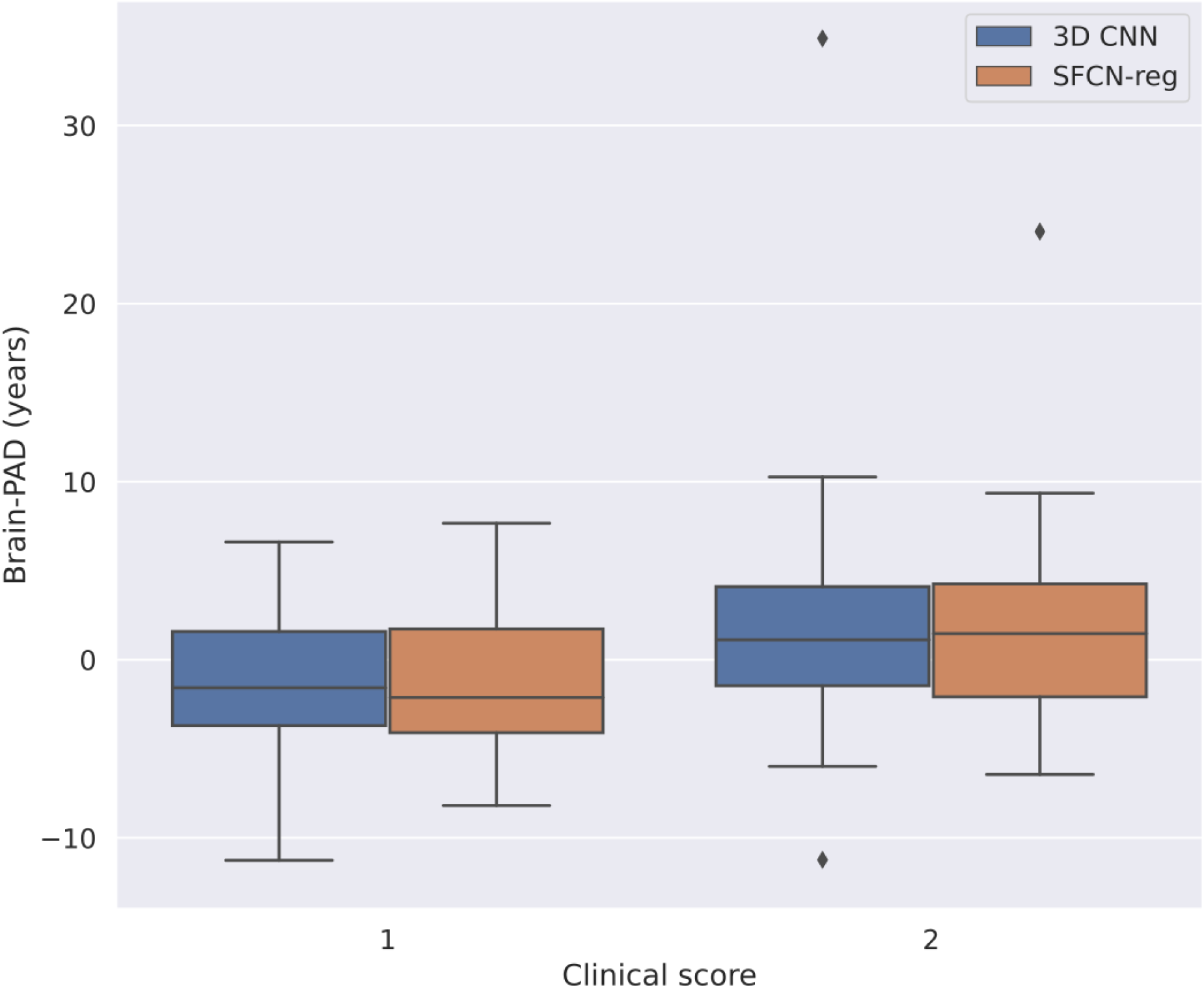
Distribution of brain-predicted age difference (brain-PAD) for all MR-ART subjects (N = 54) with a good (score 1) - medium (score 2) quality image pair (IS2). Brain-PAD is defined as brain-predicted minus chronological age in years.

**Table 6.**
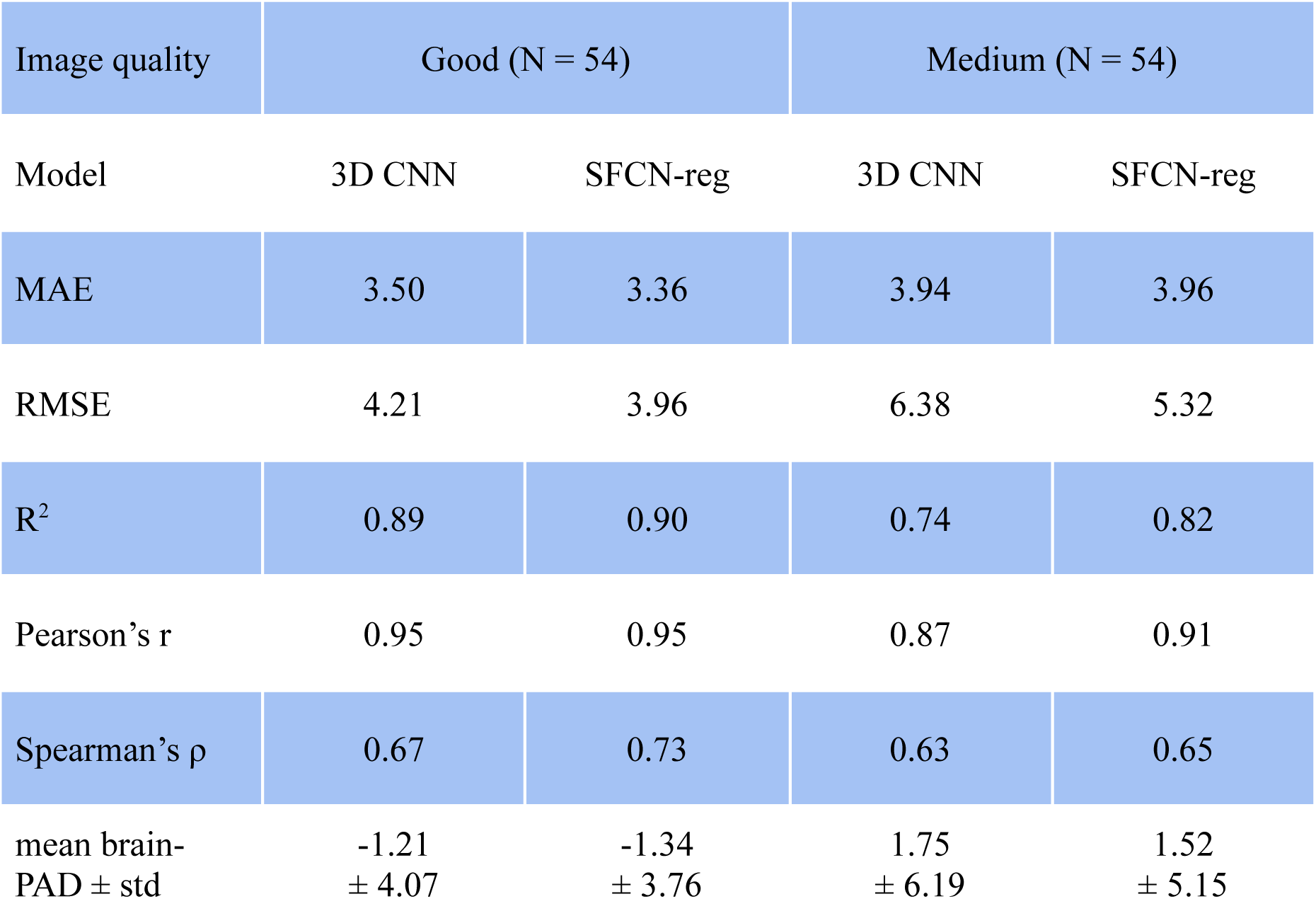
Brain age prediction performance for all MR-ART subjects (N = 54) with a good-medium quality image pair (IS2). All correlations are significant with p < 0.001. MAE = mean absolute error (in years), RMSE = root mean squared error (in years), R^2^ = coefficient of determination, brain-PAD = brain-predicted age difference (predicted minus chronological age in years), std = standard deviation.

The above reported findings were robust in that we obtained the same results after performing either of the two brain age bias correction procedures (section S1.2 in Supplementary materials and methods).

### 3.3. Correlation between brain-predicted age difference and image quality metrics

By considering all available MR-ART images, significant correlations (p_c_ < 0.05) were found between brain-PAD and all IQMs for both 3D CNN and SFCN-reg models regardless of the type of correlation coefficient (Pearson and Spearman, Fig. 9. top row). The sign of all correlations suggested increase of brain-PAD for decrease of image quality. The absolute value of correlation coefficients (r_abs_) was in the [0.18 0.68] range indicating strongest correlations between brain-PAD and cjv, cnr, snr_gm, snr_wm, snr_total and Euler IQMs. When splitting the images according to their clinical image quality score, a very similar profile of correlation coefficients was found for bad quality images (Score 3, Fig. 9. bottom row), but with slightly weaker correlations in the r_abs_ = [0.25 0.66] range and lack of significant correlations for the snr_csf IQM. For medium quality images (Score 2), the profile of correlation coefficient was similar to that found for bad quality images, but with much less significant correlations and lower absolute coefficient values (r_abs_ = [0.20 0.47]). In the case of good quality images (Score 1), the level of association further decreased resulting in significant correlations found only for the cjv IQM (r_abs_ = [0.26 0.27]). Compared to the correlations obtained by splitting the images according to their clinical image quality score (Fig. 9) a very similar profile of correlations was found when splitting the images based on their experimental conditions (STAND, HM1 and HM2; see Supplementary Fig. 3). However, the number and the strength of significant correlations were greater when splitting the images according to their experimental condition.

**Fig. 9.**
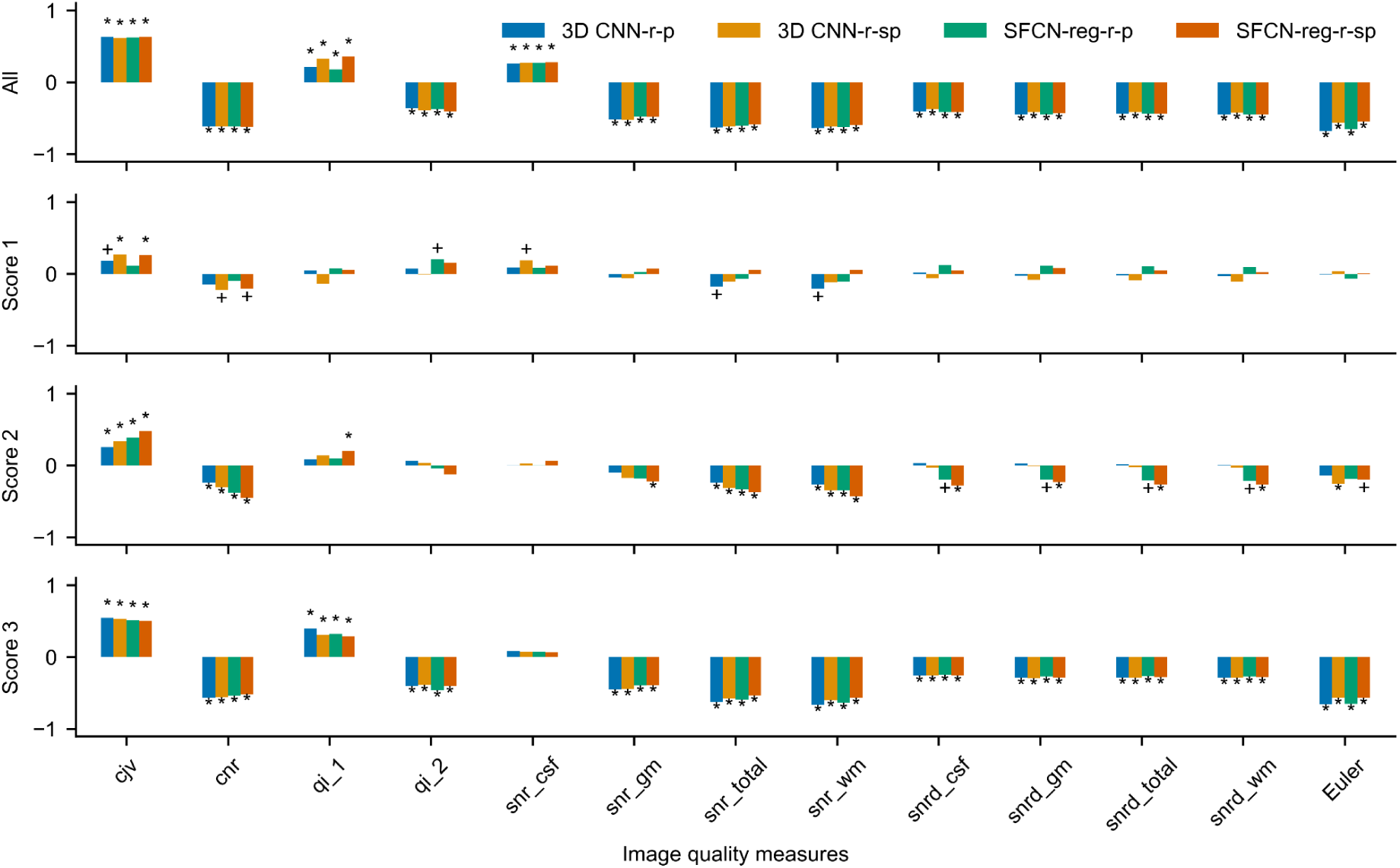
Correlations between brain predicted age difference (brain-PAD) and image quality measures for all available MR-ART images. Pearson’s (r-p) and Spearman’s (r-sp) partial correlations are shown for both 3D CNN and SFCN-reg models. Correlations were calculated by considering all images together (All) as well as by taking into account images with different clinical image quality scores (1, 2 and 3) separately. Plus and asterisk denote significant correlation coefficients without (p_u_ < 0.05) and with FDR correction (p_c_ < 0.05), respectively.

The correlation results shown in Fig. 9 were also validated by considering the IS1 selection of images assuring this way the independence of samples (Fig. 10). In general, the profile of correlations between brain-PAD and IQMs using this subsample of images was very similar to that found based on all available MR-ART scans, but there were less significant correlations. The uncorrected significant correlations (p_u_) found for Score 2 images did not survive the FDR correction, and significant correlation was found for Score 1 images only in the case of the SFCN-reg model. The strength of significant correlations obtained on selected images (all images r_abs_ = [0.21 0.66]; Score 1 r_abs_ = [0.46 0.50], Score 3 r_abs_ = [0.37 0.68]) was comparable to those found based on all available MR-ART scans. Testing the correlations between corrected brain-PAD measures and the IQMs on the selected images revealed a very similar distribution of correlation coefficients (Supplementary Figs. 4 and 5) to those obtained using the uncorrected brain-PAD values.

**Fig. 10.**
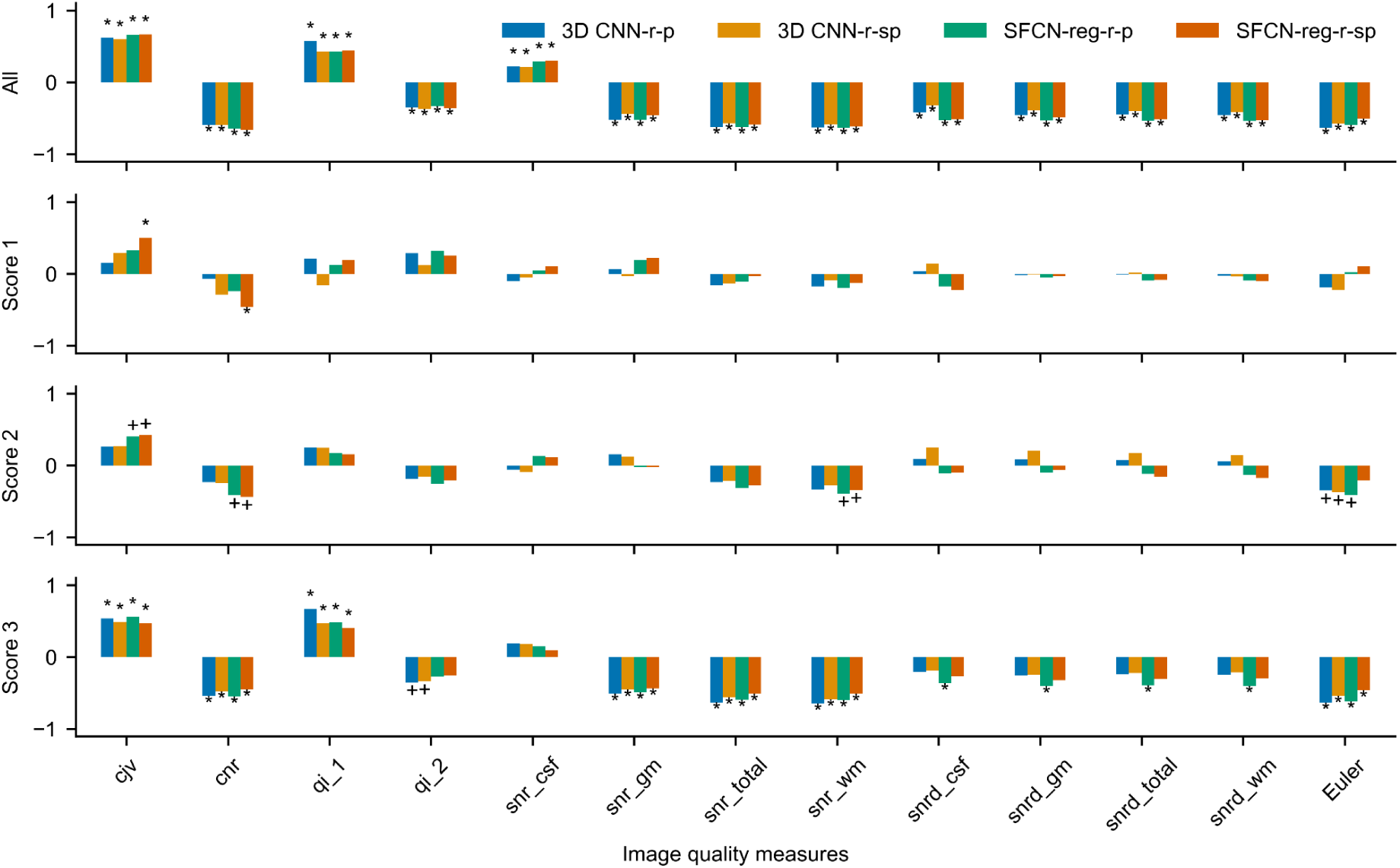
Correlations between brain predicted age difference (brain-PAD) and image quality measures for the selected MR-ART test set subjects (N = 35) with clinically good (Score 1), medium (Score 2), and bad quality (Score 3) images acquired without (STAND), with low (HM1), and with high (HM2) levels of head motion, respectively (IS1). Pearson’s (r-p) and Spearman’s (r-sp) partial correlations are shown for both 3D CNN and SFCN-reg models. Correlations were calculated by considering the images of selected subjects all together (All) as well as by taking into account images with different clinical image quality scores (1, 2 and 3) separately. Plus and asterisk denote significant correlation coefficients without (p_u_ < 0.05) and with FDR correction (p_c_ < 0.05), respectively.

Finally, to investigate the background of differences that could be observed between Pearson’s and Spearman’s correlation coefficients, we also inspected the scattergrams between the brain-PAD and the IQMs showing the strongest correlations (Figs. 11 and 12). Although the scattergrams indicate linear relationship between the variables in general, weak nonlinearity could also be observed for some of the IQMs depending also on the image quality. No substantial differences could be found between the 3D CNN and SFCN-reg models considering the scattergrams, and the correction of brain-predicted age does not seem to affect the scattergrams either.

**Fig. 11.**
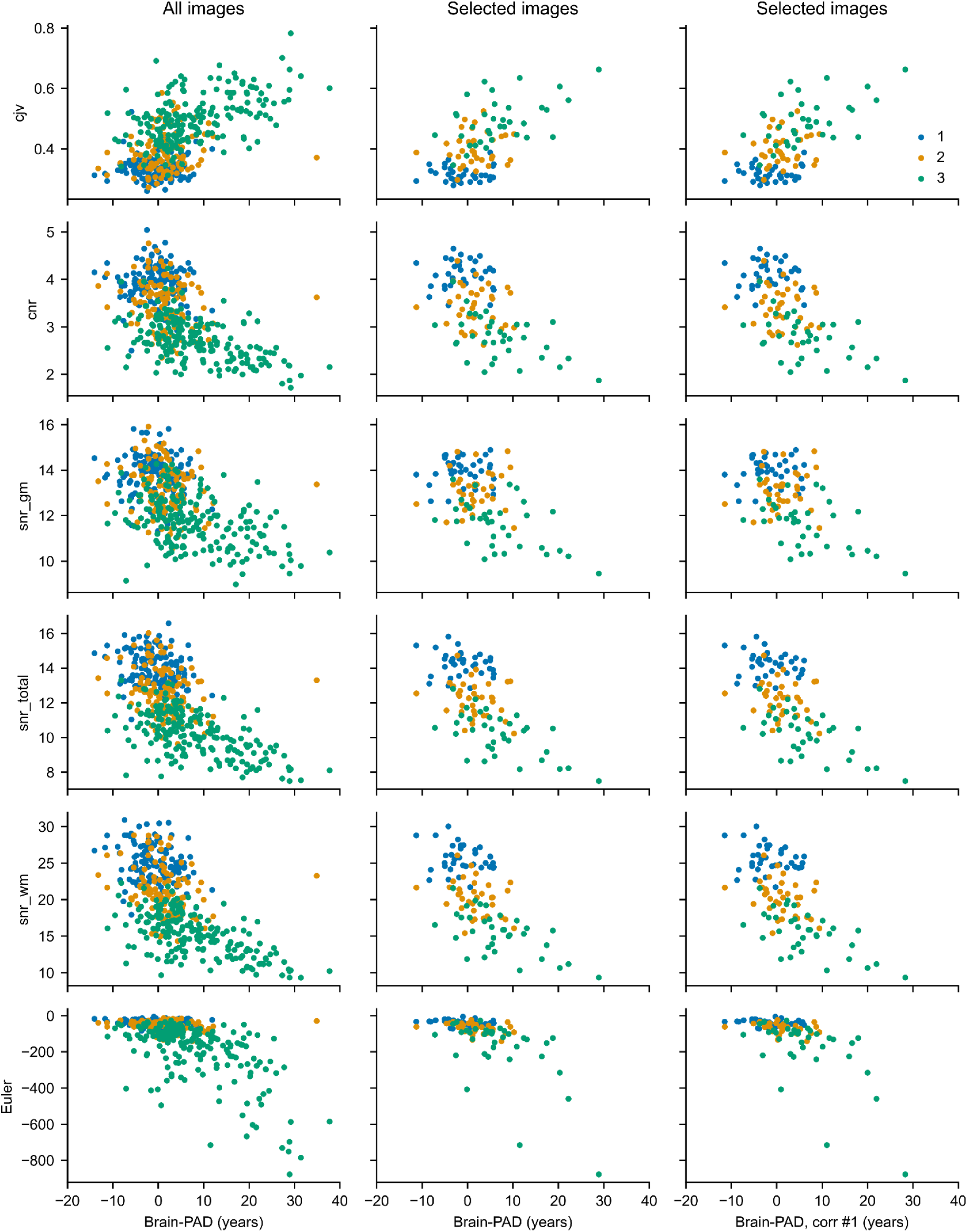
Scattergrams between brain predicted age difference (brain-PAD) and image quality measures for the 3D CNN model. Scatter plots were generated based on all available MR-ART images (left column) and by considering the selected MR-ART test set subjects (N = 35, middle and right columns) with clinically good (Score 1, blue), medium (Score 2, ocher), and bad quality (Score 3, green) images acquired without (STAND), with low (HM1), and with high (HM2) levels of head motion, respectively (IS1). In the right column, brain-PAD with correction #1 was used (see section 1.2 in the Supplementary Material).

**Fig. 12.**
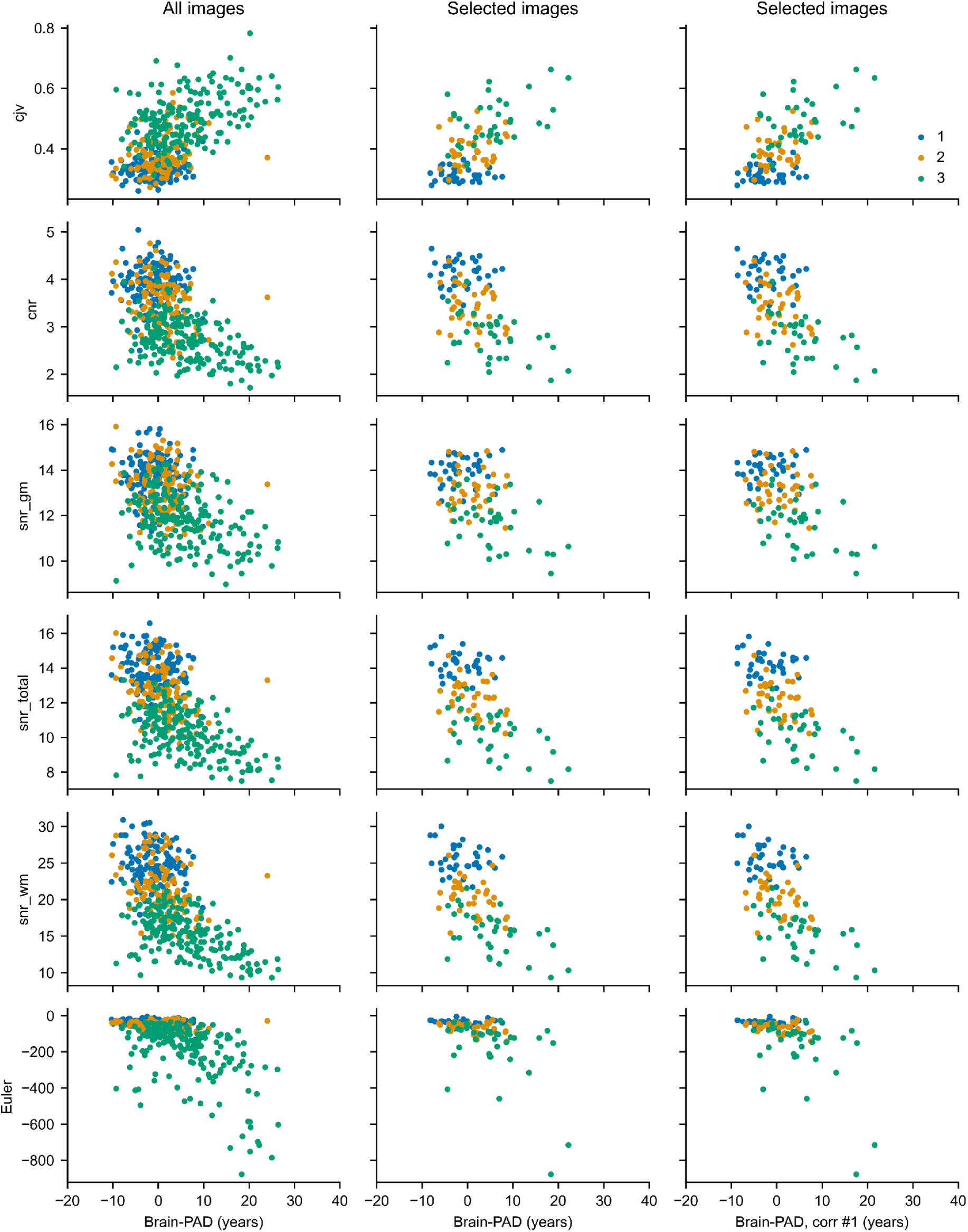
Scattergrams between brain predicted age difference (brain-PAD) and image quality measures for the SFCN-reg model. Scatter plots were generated based on all available MR-ART images (left column) and by considering the selected MR-ART test set subjects (N = 35, middle and right columns) with clinically good (Score 1, blue), medium (Score 2, ocher), and bad quality (Score 3, green) images acquired without (STAND), with low (HM1), and with high (HM2) levels of head motion, respectively (IS1). In the right column, brain-PAD with correction #1 was used (see section 1.2 in the Supplementary Material).

## 4. Discussion

In the present work, we have shown that the degradation of image quality due to head motion produces a systematic bias in brain-predicted age based on structural MRI scans. In particular, we trained two different 3D convolutional neural network models using a large amount of data to predict the age of participants based on minimally preprocessed T1-weighted scans and observed that head motion increases brain-PAD – the estimated age of the participants increased relative to their chronological age as the quality of the brain scans deteriorated. Importantly, this effect remained significant even when excluding bad quality scans that were deemed unusable from a clinical diagnostic perspective. Moreover, we found significant correlations between brain-PAD and several different IQMs, showing in general that worsening image quality was associated with increased brain-PAD. This association was the strongest for bad quality (score 3) images and the weakest for good quality (score 1) ones, however, even in the latter case a significant correlation was found between brain-PAD and the cjv IQM related to head motion artifacts [58]. The results were robust when applying previously established bias correction methods to brain-predicted age. Taken together, these findings suggest that head motion is a potential confounding factor when using brain-predicted age based on structural neuroimaging data as a proxy measure for brain health.

The systematic effect of head-motion on brain-PAD demonstrated in the current study is especially problematic because it mimics the effect of advanced brain aging. Thus, increased brain age reported in certain neuropsychiatric and neurodegenerative disease states may be confounded by the effects of head motion on image quality. For example, several studies have provided evidence for a higher brain-PAD in schizophrenia, mild cognitive impairment, and Alzheimer’s disease than in healthy controls (for reviews, see [74,75]). Given the known propensity for movement during MRI scanning in these patient populations [45,47], some of the reported effects may be overestimated when head motion is not rigorously controlled for. Moreover, given that motion may potentially be correlated with the severity of symptoms and medication use, extra care should be taken when brain-PAD is applied for the assessment of disease severity and the evaluation of treatment outcomes.

Due to the black box nature of deep neural network algorithms [76,77], further research is required to uncover the source of the observed bias in brain-predicted age. It is possible that head motion introduces artifacts to the image that mimic the signs of advanced brain aging. Previous research has shown that motion has a systematic effect on brain morphometric measures derived from T1-weighted MRI scans. In these studies, head motion was associated with a decrease in cortical gray matter thickness and volume estimates [45,51,78,79]. This bias, observed when using several different image processing software packages, does not seem to reflect the processing failure of these tools, but rather stem from alterations in the images themselves that resemble the loss of brain tissue [51]. Moreover, in one study, a substantial overlap was reported between the brain regions most affected by movement-related effects and areas that are reported to show pronounced age-related cortical atrophy [79]. These findings have potential implications for brain age prediction as well. A recent study has shown that the predictions of an XGBoost brain age algorithm [52], which uses the measurements output by the popular morphometric tool FreeSurfer [72] as features, had a modest correlation with image quality [81]. In the present work, the neural network models were trained end-to-end to predict age based on minimally preprocessed structural images rather than precomputed morphometric estimates. However, their particularly good performance on the task might be the result of successfully learning to extract features from the image that, on the one hand, reflect age-related neuroanatomical changes, and on the other hand may be systematically biased by motion-induced artifacts that mimic brain atrophy.

Several different methods have been applied to tackle the issue of head motion during MR image acquisition, however, no universal solution exists to this problem. Apart from motion restriction, shortening scanning duration may attenuate movement-related artifacts, as head motion has been shown to increase over time in the scanner, and this effect is amplified in older age [82]. Extremely rapid T1-weighted brain scans (∼1 min scanning time) have been demonstrated to produce reliable morphometric measures in healthy older adults and in individuals suffering from neurodegenerative disease [83], providing a potential avenue for improving the robustness of structural brain age prediction in populations that are more prone to motion. Visual assessment by experts can help alleviate the movement-induced reduction in the estimates provided by morphometric analysis tools, however, evidence shows that this effect is detectable even when analyzing only those images that passed a rigorous quality checking procedure and are free from visible motion artifacts [45,51,78]. Similarly, we found a significant association between brain-PAD and the cjv IQM, a metric that is sensitive to head motion artifacts [58], even in the case of clinically good quality images. This underlines the importance of controlling for movement even when the analysis is restricted to only good quality images. Including a measure of head motion—derived from an online motion-tracking system [51], from functional MRI measurements acquired in the same session [45,78,79], or from retrospective quality assessment by an image processing software tool [84]—as a covariate in the statistical model may help further mitigate the potential biasing effect of movement in brain age estimation. Although we observed an overall linear relationship between brain-PAD and IQMs, weak deviations from this pattern were also apparent, suggesting that non-linear correction methods may be even more effective in correcting the aforementioned bias. In addition, several methods are available for the correction of motion-corrupted images—e.g. prospective motion correction using volumetric navigators (vNavs) can be applied effectively to reduce the motion-induced bias in morphometric estimates [85], and recent deep learning-based algorithms have provided promising results in the retrospective removal of motion artifacts from MRI scans [86,87], although the potential benefit of these algorithms in the context of brain age prediction remains to be investigated.

Despite their complexity, 3D convolutional neural networks have shown exceptional performance in various domains of medical image analysis [88], and can be used to provide accurate and biologically relevant age prediction based on structural MRI scans [19–23]. Recently, there is an abundance of different neural network architectures developed for brain age prediction. Our results suggest that, besides optimizing to estimate individual age and testing for the association between brain-PAD and different biological and clinical phenotypes, increasing the robustness of the models to the degradation of image quality should also be of high priority in model development. 2D CNN architectures commonly used for the recognition of natural images are known to be susceptible to image distortions including but not limited to blur [89–94]. Training the networks on degraded images can improve classification performance [92,94–96], however, these models might generalize poorly to types of distortions they have not encountered previously [94,95]. Accordingly, using an ensemble of expert networks specialized to different types of distortions has been shown to increase the robustness of natural image recognition to low quality images [95,97]. Moreover, certain modifications comprising the addition of extra layers to existing architectures can increase the inherent robustness of the networks to image degradation [91,98]. Brain age prediction using deep convolutional neural networks may also benefit from the application of these strategies to tackle the issue of head motion and potentially other sources of deteriorated image quality.

A limitation of the current study is that the MR-ART test set covers only a part of the human lifespan. The relationship between head motion and brain-PAD should be investigated in younger children and adolescents as well, given the increased tendency for movement in these participant groups [45]. In the case of MR-ART subjects, the correlations between brain-PAD and IQMs sensitive to movement were observed despite controlling for the potential covariate effects of chronological age, suggesting that these associations are rather stable during adulthood. A further limitation is that the brain age prediction models in the present study were trained solely on good quality research-grade T1-weighted scans. It is likely that using training and validation sets that are more heterogeneous in terms of image quality would yield models that generalize better to artifactual MRI scans. However, severely motion-corrupted images are rarely archived and shared either in research projects or in clinical settings, making it difficult to compile a sufficiently large and qualitatively diverse dataset for developing robust deep neural networks. An alternative strategy is data augmentation using motion artifact simulation, however, even in this case it is essential to test the algorithms on real-world imaging data.

## 5. Conclusions

Our results demonstrate that brain age prediction is susceptible to movement-induced artifacts, and that head motion may result in a spurious effect of advanced brain aging even in scans deemed usable from a clinical diagnostic point of view. Special care must be taken when using brain-predicted age as a proxy for brain health, including the use of image quality assessment and control protocols and the development of robust algorithms for brain age prediction.

## Data Availability

The MR-ART dataset is publicly available, the UK Biobank dataset is available by application, and the transfer learning dataset is available upon reasonable request to the authors.

## Acknowledgements

This work was supported by Project no. RRF-2.3.1-21-2022-00009 and RRF-2.3.1-21-2022-00011, which have been implemented with the support provided by the European Union, and by a grant from the Hungarian National Research, Development and Innovation Office (2019-2.1.7-ERA-NET-2020-00008) to Z.V.. Author B.W. was supported by the Consolidator Researcher program of the Óbuda University. This research has been conducted using the UK Biobank Resource under Application Number 27236.

