## Supplementary Material for "The effect of head motion on brain age prediction using deep convolutional neural networks"

### 1. Supplementary materials and methods

#### 1.1. Details of the neural network architectures

Supplementary Table 1. Architecture of the 3D-CNN model used for brain age prediction. 3D convolutional layers are specified using the format 'CONV3D(I×J×K@F)', where I, J, and K denote the depth, height, and width of the 3D convolution window, respectively, and F denotes the number of filters. Similarly, the I, J, and K in 'MAXPOOL(I×J×K)' denote the dimensions of the max pooling window. The dropout layer applies a dropout rate of 0.4 during training. BN = batch normalization; ReLU = rectified linear unit; GAP = global average pooling.

| **Layer name** | **Layer type** | **Stride** | **Padding** | **Output shape** |
|---|---|---|---|---|
| input | Input | - | - | 182×218×182×1 |
| conv_block_1/conv1_1 | CONV3D(5×1×1@16) | 2×1×1 | SAME | 91×218×182×16 |
| conv_block_1/conv1_2 | CONV3D(1×5×1@16) | 1×2×1 | SAME | 91×109×182×16 |
| conv_block_1/conv1_3 | CONV3D(1×1×5@16) | 1×1×2 | SAME | 91×109×91×16 |
| conv_block_1/bn_1 | BN | - | - | 91×109×91×16 |
| conv_block_1/relu_1 | Activation (ReLU) | - | - | 91×109×91×16 |
| conv_block_1/conv2_1 | CONV3D(5×1×1@32) | 1×1×1 | SAME | 91×109×91×32 |
| conv_block_1/conv2_2 | CONV3D(1×5×1@32) | 1×1×1 | SAME | 91×109×91×32 |
| conv_block_1/conv2_3 | CONV3D(1×1×5@32) | 1×1×1 | SAME | 91×109×91×32 |
| conv_block_1/bn_2 | BN | - | - | 91×109×91×32 |
| conv_block_1/relu_2 | Activation (ReLU) | - | - | 91×109×91×32 |
| max_pool_1 | MAXPOOL(3×3×3) | 2×2×2 | VALID | 45×54×45×32 |
| conv_block_2/conv1_1 | CONV3D(3×1×1@32) | 1×1×1 | SAME | 45×54×45×32 |
| conv_block_2/conv1_2 | CONV3D(1×3×1@32) | 1×1×1 | SAME | 45×54×45×32 |
| conv_block_2/conv1_3 | CONV3D(1×1×3@32) | 1×1×1 | SAME | 45×54×45×32 |
| conv_block_2/bn_1 | BN | - | - | 45×54×45×32 |
| conv_block_2/relu_1 | Activation (ReLU) | - | - | 45×54×45×32 |
| conv_block_2/conv2_1 | CONV3D(3×1×1@64) | 1×1×1 | SAME | 45×54×45×64 |
| conv_block_2/conv2_2 | CONV3D(1×3×1@64) | 1×1×1 | SAME | 45×54×45×64 |

| | | | | |
|---|---|---|---|---|
| conv_block_2/conv2_3 | CONV3D(1×1×3@64) | 1×1×1 | SAME | 45×54×45×64 |
| conv_block_2/bn_2 | BN | - | - | 45×54×45×64 |
| conv_block_2/relu_2 | Activation (ReLU) | - | - | 45×54×45×64 |
| max_pool_2 | MAXPOOL(3×3×3) | 2×2×2 | VALID | 22×26×22×64 |
| conv_block_3/conv1_1 | CONV3D(3×1×1@64) | 1×1×1 | SAME | 22×26×22×64 |
| conv_block_3/conv1_2 | CONV3D(1×3×1@64) | 1×1×1 | SAME | 22×26×22×64 |
| conv_block_3/conv1_3 | CONV3D(1×1×3@64) | 1×1×1 | SAME | 22×26×22×64 |
| conv_block_3/bn_1 | BN | - | - | 22×26×22×64 |
| conv_block_3/relu_1 | Activation (ReLU) | - | - | 22×26×22×64 |
| conv_block_3/conv2_1 | CONV3D(3×1×1@128) | 1×1×1 | SAME | 22×26×22×128 |
| conv_block_3/conv2_2 | CONV3D(1×3×1@128) | 1×1×1 | SAME | 22×26×22×128 |
| conv_block_3/conv2_3 | CONV3D(1×1×3@128) | 1×1×1 | SAME | 22×26×22×128 |
| conv_block_3/bn_2 | BN | - | - | 22×26×22×128 |
| conv_block_3/relu_2 | Activation (ReLU) | - | - | 22×26×22×128 |
| max_pool_3 | MAXPOOL(3×3×3) | 2×2×2 | VALID | 10×12×10×128 |
| conv_block_4/conv1_1 | CONV3D(3×1×1@128) | 1×1×1 | SAME | 10×12×10×128 |
| conv_block_4/conv1_2 | CONV3D(1×3×1@128) | 1×1×1 | SAME | 10×12×10×128 |
| conv_block_4/conv1_3 | CONV3D(1×1×3@128) | 1×1×1 | SAME | 10×12×10×128 |
| conv_block_4/bn_1 | BN | - | - | 10×12×10×128 |
| conv_block_4/relu_1 | Activation (ReLU) | - | - | 10×12×10×128 |
| conv_block_4/conv2_1 | CONV3D(3×1×1@256) | 1×1×1 | SAME | 10×12×10×256 |
| conv_block_4/conv2_2 | CONV3D(1×3×1@256) | 1×1×1 | SAME | 10×12×10×256 |
| conv_block_4/conv2_3 | CONV3D(1×1×3@256) | 1×1×1 | SAME | 10×12×10×256 |
| conv_block_4/bn_2 | BN | - | - | 10×12×10×256 |
| conv_block_4/relu_2 | Activation (ReLU) | - | - | 10×12×10×256 |
| avg_pool | GAP | - | - | 256 |
| dense_hidden | Dense (fully connected) | - | - | 128 |

| | | | | |
|---|---|---|---|---|
| dense_hidden_relu | Activation (ReLU) | - | - | 128 |
| dropout | Dropout (0.4) | - | - | 128 |
| output | Dense (fully connected) | - | - | 1 |

Supplementary Table 2. Architecture of the SFCN-reg model used for brain age prediction. 3D convolutional layers are specified using the format 'CONV3D(I×J×K@F)', where I, J, and K denote the depth, height, and width of the 3D convolution window, respectively, and F denotes the number of filters. Similarly, the I, J, and K in 'MAXPOOL(I×J×K)' denote the dimensions of the max pooling window. The dropout layer applies a dropout rate of 0.5 during training. BN = batch normalization; ReLU = rectified linear unit; GAP = global average pooling.

| Layer name | Layer type | Stride | Padding | Output shape |
|---|---|---|---|---|
| input | Input | - | - | 182×218×182×1 |
| block_1/conv | CONV3D(3×3×3@32) | 1×1×1 | SAME | 182×218×182×32 |
| block_1/bn | BN | - | - | 182×218×182×32 |
| block_1/max_pool | MAXPOOL(2×2×2) | 2×2×2 | VALID | 91×109×91×32 |
| block_1/relu | Activation (ReLU) | - | - | 91×109×91×32 |
| block_2/conv | CONV3D(3×3×3@64) | 1×1×1 | SAME | 91×109×91×64 |
| block_2/bn | BN | - | - | 91×109×91×64 |
| block_2/max_pool | MAXPOOL(2×2×2) | 2×2×2 | VALID | 45×54×45×64 |
| block_2/relu | Activation (ReLU) | - | - | 45×54×45×64 |
| block_3/conv | CONV3D(3×3×3@128) | 1×1×1 | SAME | 45×54×45×128 |
| block_3/bn | BN | - | - | 45×54×45×128 |
| block_3/max_pool | MAXPOOL(2×2×2) | 2×2×2 | VALID | 22×27×22×128 |
| block_3/relu | Activation (ReLU) | - | - | 22×27×22×128 |
| block_4/conv | CONV3D(3×3×3@256) | 1×1×1 | SAME | 22×27×22×256 |
| block_4/bn | BN | - | - | 22×27×22×256 |
| block_4/max_pool | MAXPOOL(2×2×2) | 2×2×2 | VALID | 11×13×11×256 |
| block_4/relu | Activation (ReLU) | - | - | 11×13×11×256 |
| block_5/conv | CONV3D(3×3×3@256) | 1×1×1 | SAME | 11×13×11×256 |
| block_5/bn | BN | - | - | 11×13×11×256 |
| block_5/max_pool | MAXPOOL(2×2×2) | 2×2×2 | VALID | 5×6×5×256 |
| block_5/relu | Activation (ReLU) | - | - | 5×6×5×256 |
| block_6/conv | CONV3D(1×1×1@64) | 1×1×1 | VALID | 5×6×5×64 |

| | | | | |
|---|---|---|---|---|
| block_6/bn | BN | - | - | 5×6×5×64 |
| block_6/relu | Activation (ReLU) | - | - | 5×6×5×64 |
| block_7/avg_pool | GAP | - | - | 64 |
| block_7/dropout | Dropout (0.5) | - | - | 64 |
| block_7/output | Dense (fully connected) | - | - | 1 |

#### 1.2. Bias correction methods for brain age prediction

In addition to the main analysis involving the original (uncorrected) model predictions, we applied two different correction methods to address the potential bias in brain age prediction consisting of a tendency to overestimate the age of younger and underestimate the age of elderly subjects, and to estimate the age of participants whose age lies closer to the mean age in the training sample more accurately [1]. Correction method #1 consisted of modeling the relationship between brain-PAD (brain-predicted age difference = brain-predicted minus chronological age) and chronological age as a first step, using the following formula:

$$brain\text{-}PAD = \alpha \times x + \beta$$

where $x$ is the chronological age. Then, predictions on the test set are corrected using the estimated slope α and intercept β:

$$\hat{y} = y - (\alpha \times x + \beta)$$

where $y$ is the original and $\hat{y}$ is the corrected brain-predicted age. This procedure was applied in [2] and corresponds to Equations (1) and (2) in [1].

Correction method #2 included modeling the relationship between brain-predicted and chronological age:

$$y = \alpha \times x + \beta$$

and then correcting brain-predicted age on the test set using the formula:

$$\hat{y} = \frac{y - \beta}{\alpha}$$

as in [3]. Importantly, we estimated the coefficients for both methods using the labels and predictions on the transfer learning validation set (similarly to [3]), and used these coefficients to correct brain-predicted age on the MR-ART test set.

#### 2. Supplementary results

Supplementary Table 3. Brain age prediction performance on the full MR-ART set (N = 148 subjects) in the standard setting without motion (STAND), and at low (HM1) and high (HM2) levels of head motion. All correlations are significant with $p < 0.001$. MAE = mean absolute error (in years), RMSE = root mean squared error (in years), $R^2$ = coefficient of determination, brain-PAD = brain-predicted age difference (brain-predicted minus chronological age in years), std = standard deviation.

| Acquisition condition | STAND (N = 148) | | HM1 (N = 141) | | HM2 (N = 147) | |
|---|---|---|---|---|---|---|
| Model | 3D CNN | SFCN-reg | 3D CNN | SFCN-reg | 3D CNN | SFCN-reg |
| MAE | 3.41 | 3.06 | 5.99 | 4.97 | 9.45 | 7.59 |
| RMSE | 4.30 | 3.87 | 9.01 | 6.95 | 12.59 | 9.99 |
| $R^2$ | 0.89 | 0.91 | 0.34 | 0.61 | 0.02 | 0.38 |
| Pearson's r | 0.94 | 0.96 | 0.74 | 0.83 | 0.73 | 0.80 |
| Spearman's ρ | 0.70 | 0.77 | 0.51 | 0.52 | 0.55 | 0.54 |
| mean brain-PAD ± std | -0.80 ± 4.24 | -1.04 ± 3.74 | 3.94 ± 8.13 | 2.98 ± 6.30 | 8.38 ± 9.43 | 6.32 ± 7.77 |

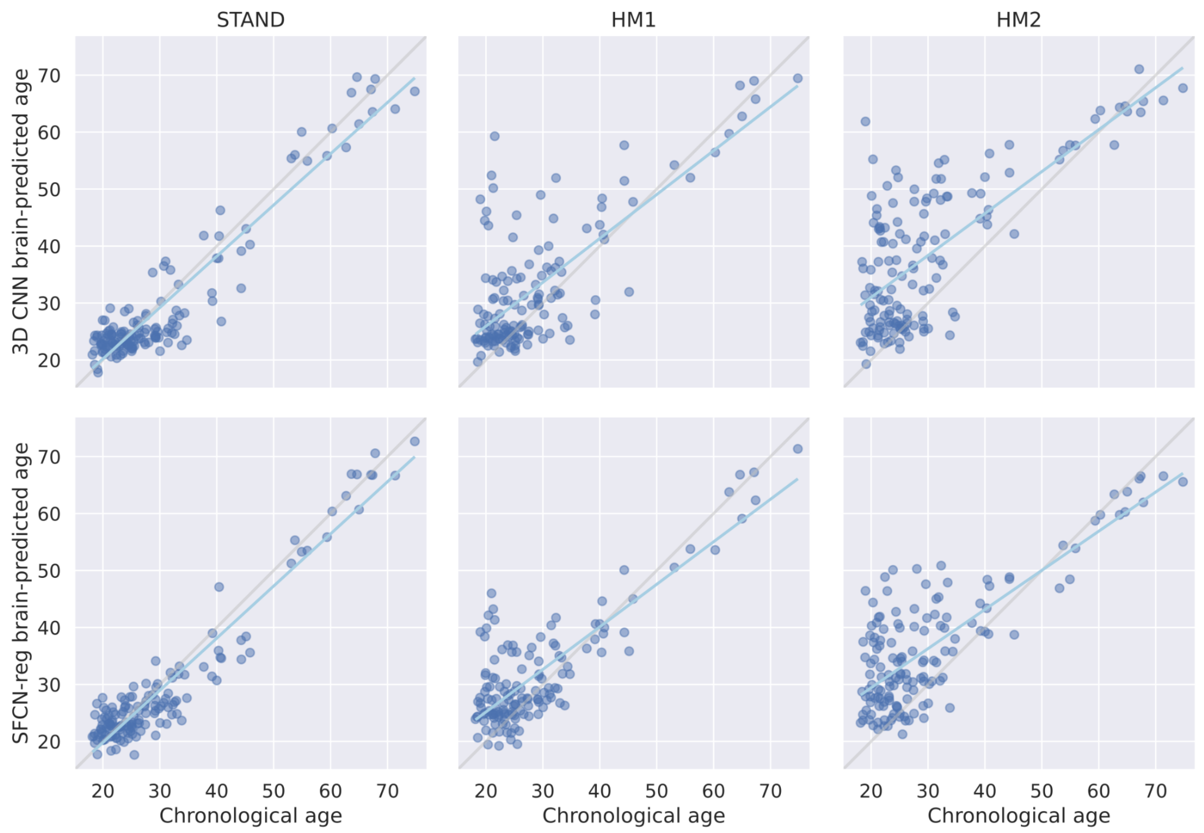


Supplementary Fig. 1. Correlations between chronological and brain-predicted age (in years) for the 3D CNN (upper row) and SFCN-reg (lower row) models on the full MR-ART set (N = 148 subjects) grouped according to acquisition condition. Gray and cyan lines correspond to the lines of identity and the regression lines, respectively. STAND = standard setting without motion; HM1 = low level of head motion (5 nods); HM2 = high level of head motion (10 nods).

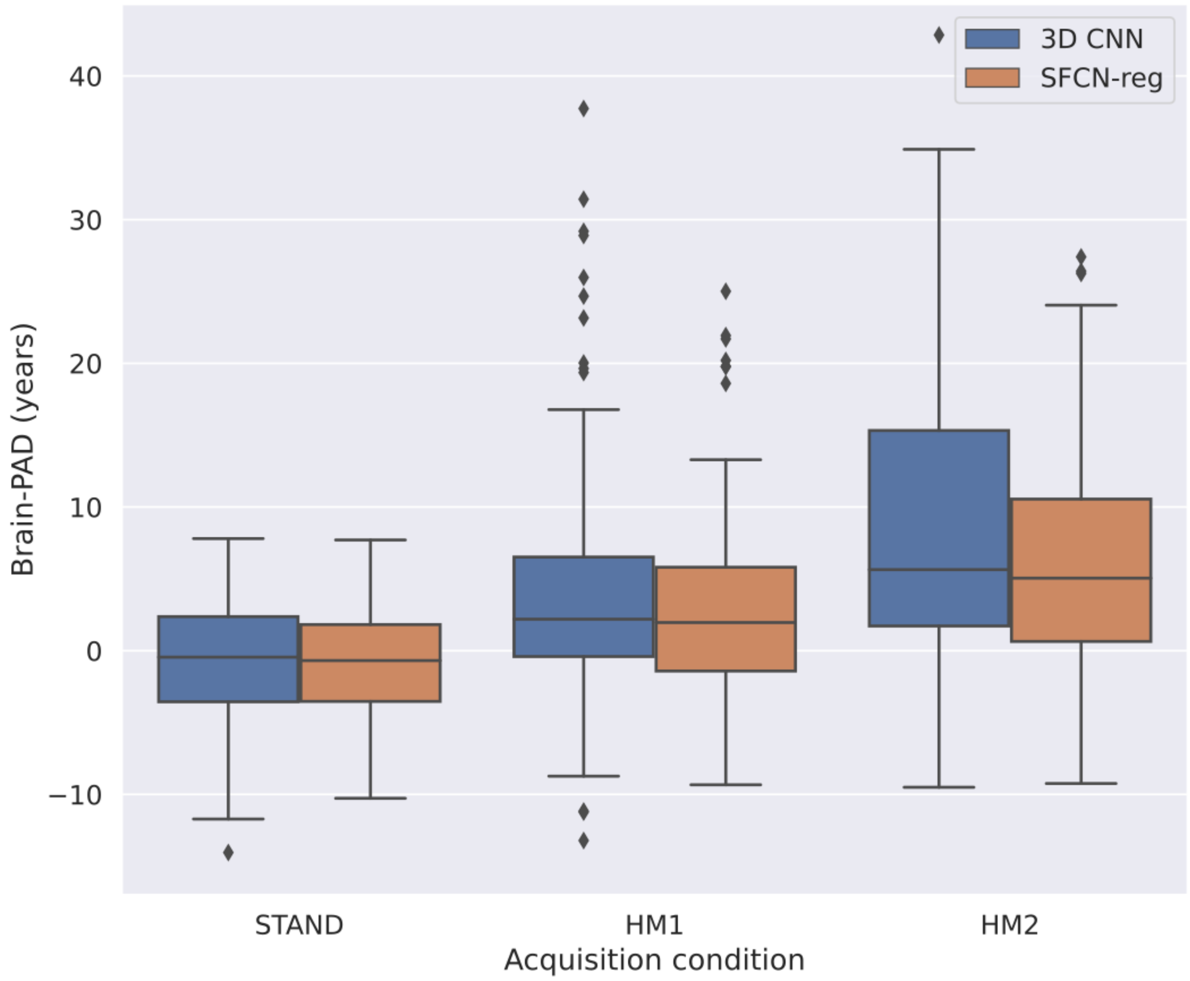


Supplementary Fig. 2. Distribution of brain-predicted age difference (brain-PAD) on the full MR-ART set (N = 148 subjects) grouped according to acquisition condition. Brain-PAD is defined as brain-predicted minus chronological age in years. STAND = standard setting without motion; HM1 = low level of head motion (5 nods); HM2 = high level of head motion (10 nods).

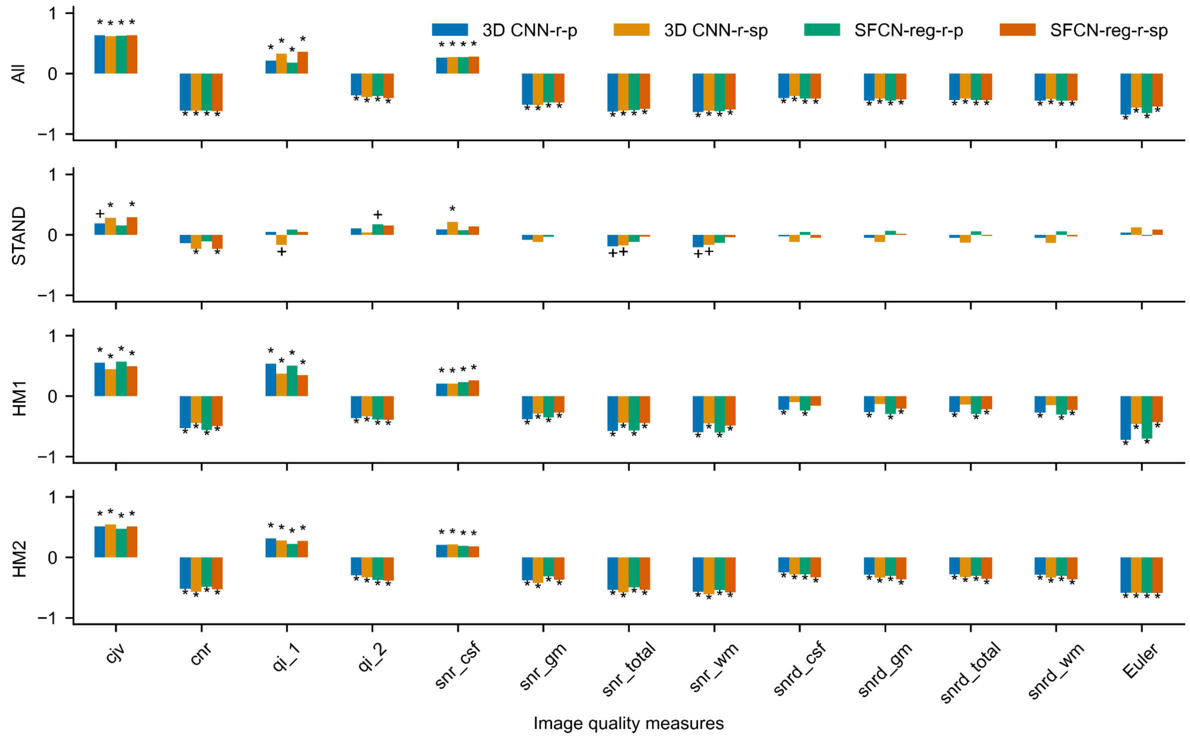


Supplementary Fig. 3. Correlations between brain-predicted age difference (brain-PAD) and image quality measures for all available MR-ART images. Pearson's (r-p) and Spearman's (r-sp) partial correlations are shown for both 3D CNN and SFCN-reg models. Correlations were calculated by considering all images together (All) as well as by taking into account images recorded in different conditions (STAND, HM1 and HM2) separately. Plus and asterisk denote significant correlation coefficients without ($p_u$ < 0.05) and with FDR correction ($p_c$ < 0.05), respectively.

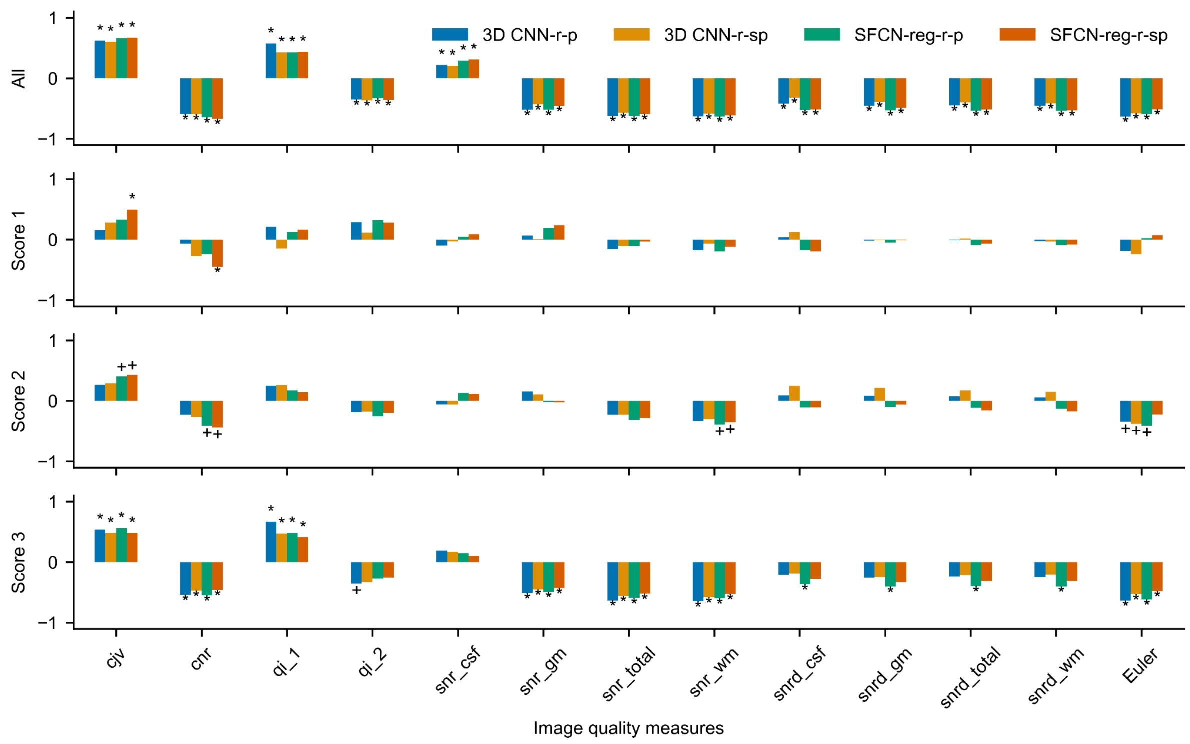


Supplementary Fig. 4. Correlations between brain predicted age difference with correction #1 (brain-PAD) and image quality measures for the selected MR-ART test set subjects (N = 35) with clinically good (Score 1), medium (Score 2), and bad quality (Score 3) images acquired without (STAND), with low (HM1), and with high (HM2) levels of head motion, respectively (IS1). Pearson's (r-p) and Spearman's (r-sp) partial correlations are shown for both 3D CNN and SFCN-reg models. Correlations were calculated by considering the images of selected subjects all together (All) as well as by taking into account images with different clinical scores (1, 2 and 3) separately. Plus and asterisk denote significant correlation coefficients without ($p_u < 0.05$) and with FDR correction ($p_c < 0.05$), respectively.

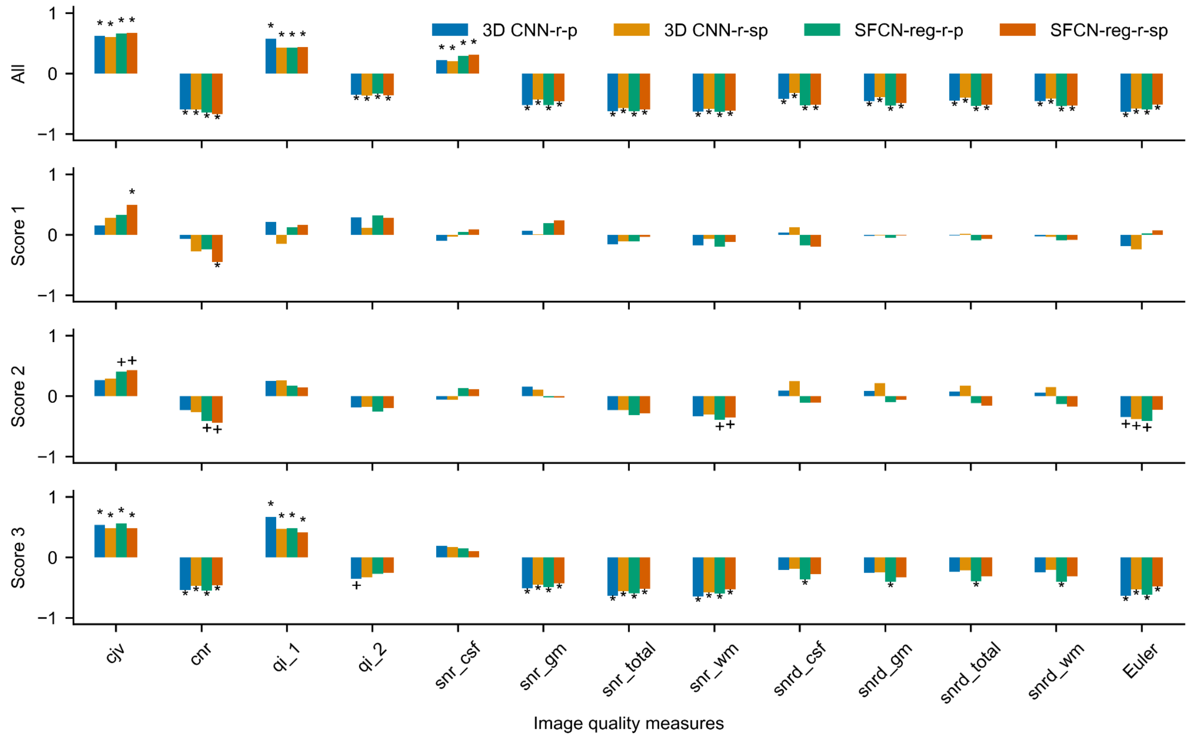


Supplementary Fig. 5. Correlations between brain predicted age difference with correction #2 (brain-PAD) and image quality measures for the selected MR-ART test set subjects (N = 35) with clinically good (Score 1), medium (Score 2), and bad quality (Score 3) images acquired without (STAND), with low (HM1), and with high (HM2) levels of head motion, respectively (IS1). Pearson's (r-p) and Spearman's (r-sp) partial correlations are shown for both 3D CNN and SFCN-reg models. Correlations were calculated by considering the images of selected subjects all together (All) as well as by taking into account images with different clinical scores (1, 2 and 3) separately. Plus and asterisk denote significant correlation coefficients without ($p_u < 0.05$) and with FDR correction ($p_c < 0.05$), respectively.
